# Open Source, Open Science: Development of OpenLESS as the Automated Landing Error Scoring System

**DOI:** 10.1101/2024.11.28.24318160

**Authors:** Jeffrey A. Turner, Elaine T. Reiche, Matthew T. Hartshorne, Connor C. Lee, Joanna M. Blodgett, Darin A. Padua

## Abstract

**Context:** The Open Landing Error Scoring System (OpenLESS) is a novel development aimed at automating the LESS for assessment of lower extremity movement quality during a jump-landing task. With increasing utilization of clinical measures to monitor outcomes and limited time during clinical visits for a lengthy analysis of functional movement, there is a pressing need to extend automation efforts. Addressing these issues, OpenLESS is an open-source tool that utilizes a freely available markerless motion capture system to automate the LESS using three-dimensional kinematics.

**Objective:** To describe the development of OpenLESS, examine the validity against expert rater LESS scores in healthy and clinically relevant cohorts, and assess the intersession reliability collected across four time points in an athlete cohort.

**Design:** Observational

**Participants:** 92 participants (72 females and 20 males, mean age 23.3 years) from healthy, post-anterior cruciate ligament reconstruction (ACLR; median 33 months since surgery), and amateur athlete cohorts.

**Main Outcome Measure(s):** A software package, “OpenLESS,” was developed to interpret movement quality (LESS score) from kinematics captured from markerless motion capture. Validity and reliability were assessed with intraclass correlation coefficients (ICC), standard error of measure (SEM), and minimal detectable change (MDC).

**Results:** OpenLESS agreed well with expert rater LESS scores for healthy (ICC_2,*k*_=0.79) and clinically relevant, post-ACLR cohorts (ICC_2,*k*_=0.88). The automated OpenLESS system reduced scoring time, processing all 159 trials in under 15 minutes compared to the 18.5 hours (7 minutes per trial) required for manual expert rater scoring. When tested outside laboratory conditions, OpenLESS showed excellent reliability across repeated sessions (ICC_2,*k*_>0.89), with a SEM of 0.98 errors and MDC of 2.72 errors.

**Conclusion:** OpenLESS shows promise as an efficient, automated tool for clinically assessing jump-landing quality, with good validity versus experts in healthy and post-ACLR populations, and excellent field reliability, addressing the need for objective movement analysis.

**KEY POINTS:**

- OpenLESS accurately detected jump-landing events (ICC>0.99) using markerless motion capture, validating its use as an alternative to laboratory-based force plate measurements.
- The automated scoring system showed good agreement with expert raters in healthy (ICC=0.79) and post-ACLR (ICC=0.88) populations.
- OpenLESS demonstrated good to excellent test-retest reliability (ICC=0.89) across multiple testing sessions, with minimal score variation, supporting its utility for longitudinal movement assessment.

---

Functional movement screening is a well-established component for assessing lower extremity injury risk in clinical and athletic populations.^1–3^ Athletic Trainers’ and Physical Therapists’ Association position statements and guidelines have recommended applying movement quality assessments to identify individuals with heightened injury risk, enabling the implementation of targeted prevention strategies such as strength training, flexibility exercises, and movement retraining.^1,2,4,5^ While laboratory-based optical 3-dimensional (3D) motion capture systems are considered the gold standard for quantifying biomechanical risk factors,^6^ their substantial time (up to 48 hours per participant) and financial (up to $150,000 per system) requirements render them impractical for large-scale injury risk screening programs.^7^ As such, there is a growing need for efficient, field-based functional assessment tools that can be readily deployed to identify high-risk movement patterns within clinical and athletic populations.

The Landing Error Scoring System (LESS) has become a widely accepted clinical tool for evaluating jump-landing mechanics.^3,5,8,9^ Initially developed to screen ‘at-risk’ individuals for non-contact injuries, video is used to capture jump landings and subsequently graded on 17 criteria over three jump-landing trials.^9^ A maximal score of 19 errors can be reached for exceptionally poor performances, with a score of <5 considered to be good (i.e., low risk). The LESS has been validated against 3D motion capture,^9–11^ and high LESS scores have been associated with movement patterns linked to increased injury risk, such as ACL injuries, including decreased flexion at the hip and knee, increased knee valgus, internal rotation moments, and elevated anterior tibial shear forces upon landing from a jump.^9,11,12^ The clinical relevance of LESS scoring is further supported by prospective research demonstrating that youth soccer athletes with higher LESS scores face greater risk of ACL injury.^5^

Building upon this foundation, Mauntel et al.^13^ subsequently developed an automated grading system for an expanded LESS version, utilizing a depth camera and Kinect sensor (Microsoft Corp, Redmond, WA) with proprietary machine learning algorithms to calculate the relevant kinematic variables. This automated system reliably estimated kinematics during drop vertical jump assessments^13^ and demonstrated moderate agreement against the gold standard 3D motion capture approach.^14^ Advancements in deep learning computer vision and markerless motion capture have enabled further efforts to automate LESS scoring.^15^ While this prior work demonstrated the feasibility of machine learning-based LESS assessment, the proprietary nature of the underlying code and methods limited broader accessibility. In contrast, open-source solutions leveraging frameworks like OpenPose^16^ and HRNet^17^ offer a more scalable path to bringing efficient, automated movement quality assessments into clinical practice.^6^

Given the growing emphasis on efficient, cost-effective assessment tools within clinical practice, automating the LESS represents a promising avenue to expand the utility and accessibility of this validated movement quality screening.^3,8^ Prior efforts to automate LESS scoring have demonstrated the technical feasibility of this approach,^13–15^ but the proprietary nature of these systems has limited their widespread adoption. In contrast, open-source frameworks leveraging markerless motion capture, such as OpenCap (Stanford University, USA),^7,18^ offer a more scalable path to integrating automated LESS assessments into clinical and athletic settings.

The primary aim of the present study was to develop and evaluate the validity and reliability of OpenLESS, an automated scoring system for the LESS utilizing open-source software and low-cost markerless motion capture (OpenCap). To demonstrate the clinical utility of this approach, OpenLESS was validated against expert rater scores in healthy and post anterior cruciate ligament reconstruction (ACLR) populations, with intersession reliability examined in an amateur athlete cohort. We hypothesized that the automated scoring software (Supplemental File 1) using markerless motion capture would be a valid and reliable version of the LESS.

## METHODS

### Design

This secondary analysis included three different cohorts from repeated measures and cross-sectional observational studies to assess the measurement properties of an automated pipeline for scoring the LESS using a portable, low-cost, markerless motion capture system. To assess validity, we compared OpenLESS scores to expert rater LESS scores in a healthy cohort and a post-ACLR cohort. Reliability of OpenLESS was assessed in a field-based athlete cohort across up to four visits over a month. This study followed the Strengthening the Reporting of Observational Studies in Epidemiology (STROBE) guidelines,^19^ ensuring comprehensive reporting and transparency (Figure 1).

**Figure 1.**
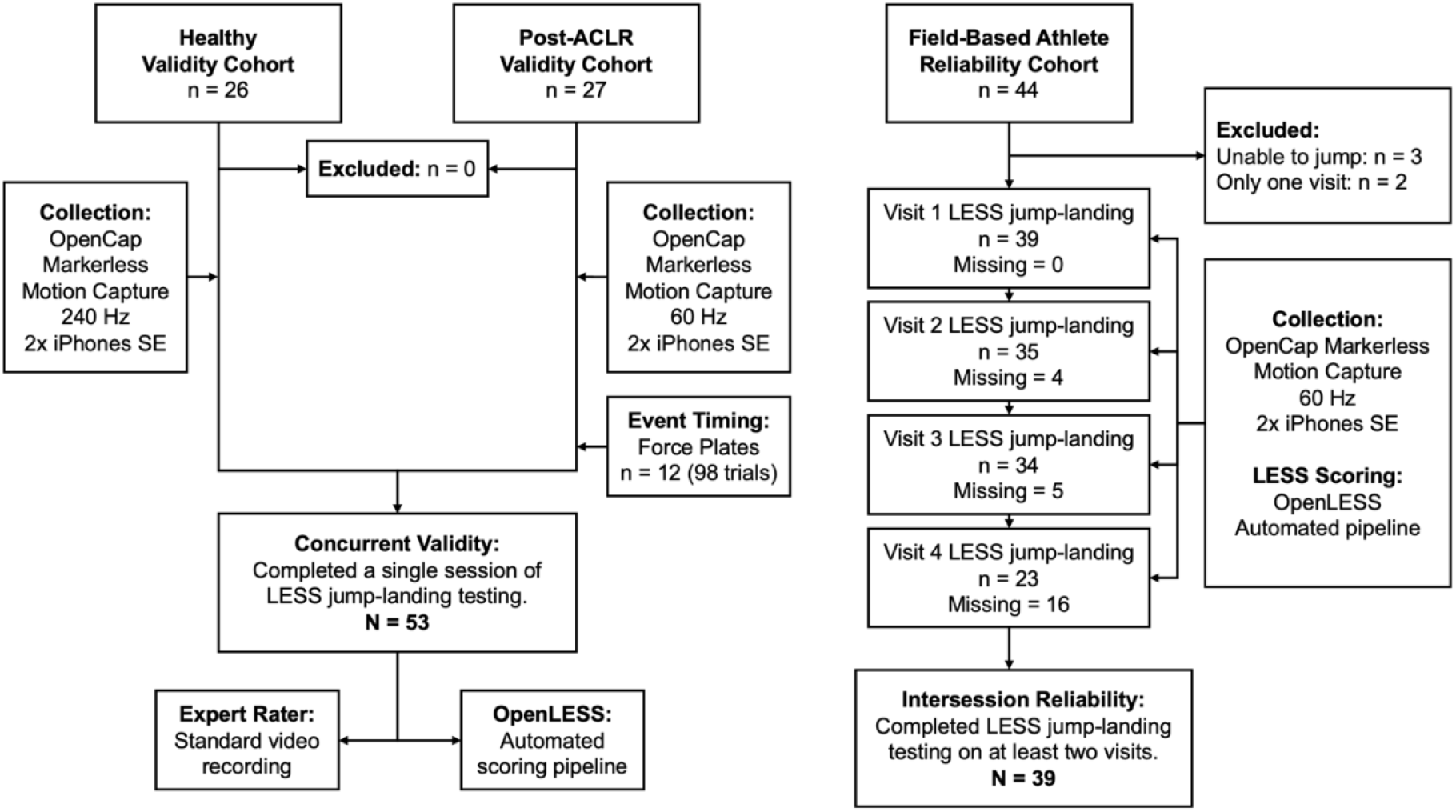
STROBE Flow Diagram.

### Participants

The healthy cohort consisted of 26 university students (12 males, 14 females) with no history of lower extremity surgery or injuries in the last 6 months, approved by the University of North Carolina’s Institutional Review Board (IRB: 23-1507). Healthy participants arrived at the biomechanics laboratory for a single session where they performed the LESS jump-landing under markerless motion capture.

The post-ACLR cohort included 27 individuals (8 males, 19 females) 6-72 months post-ACLR surgery, approved by the University of North Carolina (IRB 24-1263; 22-1599). LESS expert raters were not blinded to whether participants were healthy or post-ACLR. Post-ACLR participants arrived at the biomechanics laboratory for a single session where they performed the LESS jump-landing under markerless motion capture.

The field-based athlete cohort comprised 39 females (18 amateur soccer players, 10 university athletes from ball and non-ball sports, and 11 recreational weightlifters) with no current lower extremity injuries, approved by University College London (REC Project ID: 25759/001). All participants provided written informed consent. Athlete participants were assessed outside a laboratory environment (soccer pitch, athletic field, and indoor recreation center) for up to four consecutive sessions where they performed the LESS jump-landing under markerless motion capture. To be included, athlete participants needed to attend at least two of the four potential visits, these were allowed to be non-consecutive.

### Testing Procedures

#### The Task: LESS Jump-Landing

All cohorts’ participants performed the double-leg jump-landing rebound task under markerless motion capture, referred to as the jump-landing task (Figure 2).^9^ Participants jumped from a 30 cm box to a landing spot 50% of their height in front, landed, and then performed a maximal vertical jump. After verbal instruction up to three practice trials were allowed, and participants wore their preferred footwear and the validation cohorts were required to wear tight-fitting clothing, whereas the field-based athlete cohort were allowed to wear their usual exercise clothing.^20^ Three successful trials were then collected for use in analysis. Trial success was determined if the participant (1) jumped and landed correctly, (2) jumped vertically during the maximal jump, and (3) completed the task without losing balance.

**Figure 2.**
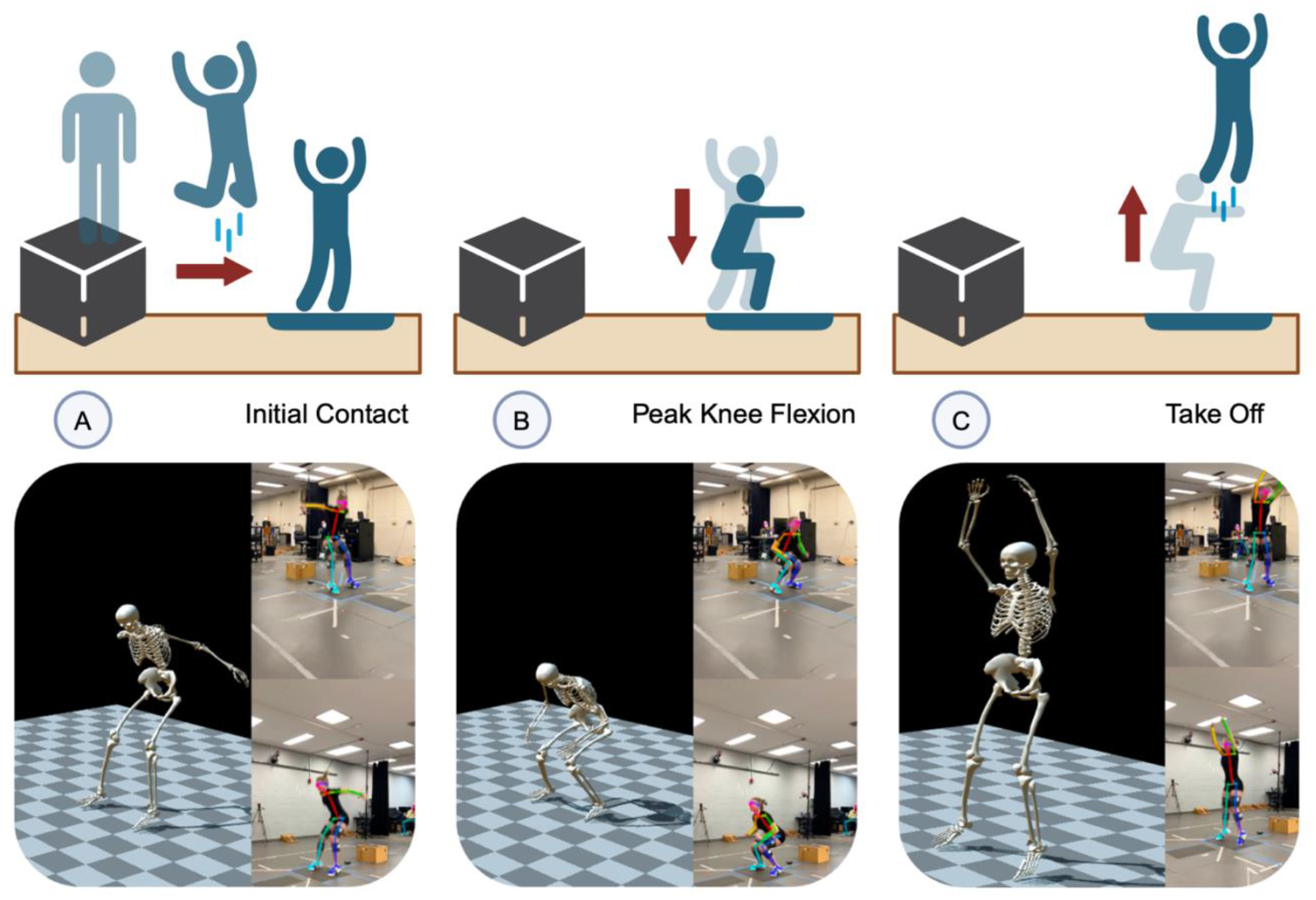
LESS Jump-Landing Task. The figure was adapted from Turner et al.^21^ with permission.

The area of interest for scoring the movement quality with the LESS was the first landing of the jump-landing task.^9,13^ The first landing was defined as the stance phase bounded by the moment of initial foot contact with the ground to take off (i.e., toe off). The stance phase was divided into a braking phase and a propulsion phase. The braking phase was defined as the time interval from the feet contacting the ground (Figure 2A) to the lowest point of the braking phase before upward movement (identified by peak knee flexion; Figure 2B). The propulsion phase was defined as the time interval from the lowest point of the braking phase before upward movement (Figure 2B) to the feet taking off the ground (Figure 2C). The lowest point of the braking phase before upward movement (Figure 2B) was considered as the transition from eccentric to concentric movement.^22^

The original LESS evaluated 17 specific movement characteristics during the jump-landing task, with items scored at initial ground contact (Figure 2A), during the braking phase (Figure 2A to 2B), and at peak knee flexion (Figure 2B).^5,9^ Each item is scored dichotomously (0 or 1) or categorically (0, 1, or 2) based on the presence or absence of movement errors where an error from either limb results in error for that item, with a total possible score ranging from 0 to 19 errors. The scoring criteria include the assessment of sagittal and frontal plane positioning of the trunk, hips, and knees, and additional items for overall movement quality and symmetry.^5,9^ The LESS has demonstrated good interrater (ICC_2,*k*_ = 0.84, SEM = 0.71) and intersession (ICC_2,*k*_ = 0.81, SEM = 0.81) reliability, with higher scores indicating more aberrant movement patterns that may increase injury risk.^9,10^ We used a modified version of the LESS, expanded to 19 items by Mauntel et al.^13^ to include two additional asymmetrical landing characteristics, who demonstrated strong agreement between an automated grading tool and expert raters.

#### The System: OpenCap Markerless Motion Capture

The 3D kinematics of the trunk and lower extremities were collected using two smartphones (iPhone 12 SE, Apple Inc., Cupertino, CA, USA) running OpenCap v0.3 (Stanford, USA) markerless motion capture software.^23^ OpenCap has been validated against optoelectronic 3D motion capture systems.^7,18,24^ The smartphones were positioned at a standardized distance of 3 meters from the landing area, 1.5 meters off the ground, and at 45° angles (Figure 3). We followed OpenCap’s best practice guidelines^7^ for smartphone setup, calibration with a 720×540 mm checkerboard, and the recording of a static trial. Markerless motion capture was sampled at 60 Hz for the post-ACLR and field-based athlete cohorts and 240 Hz for the healthy cohort using the default pose estimation algorithm (HRnet)^17^ via OpenCap’s cloud-based software (v0.3).^7^ Further technical details can be found in the development and validation paper by Uhlrich et al.^7^

**Figure 3.**
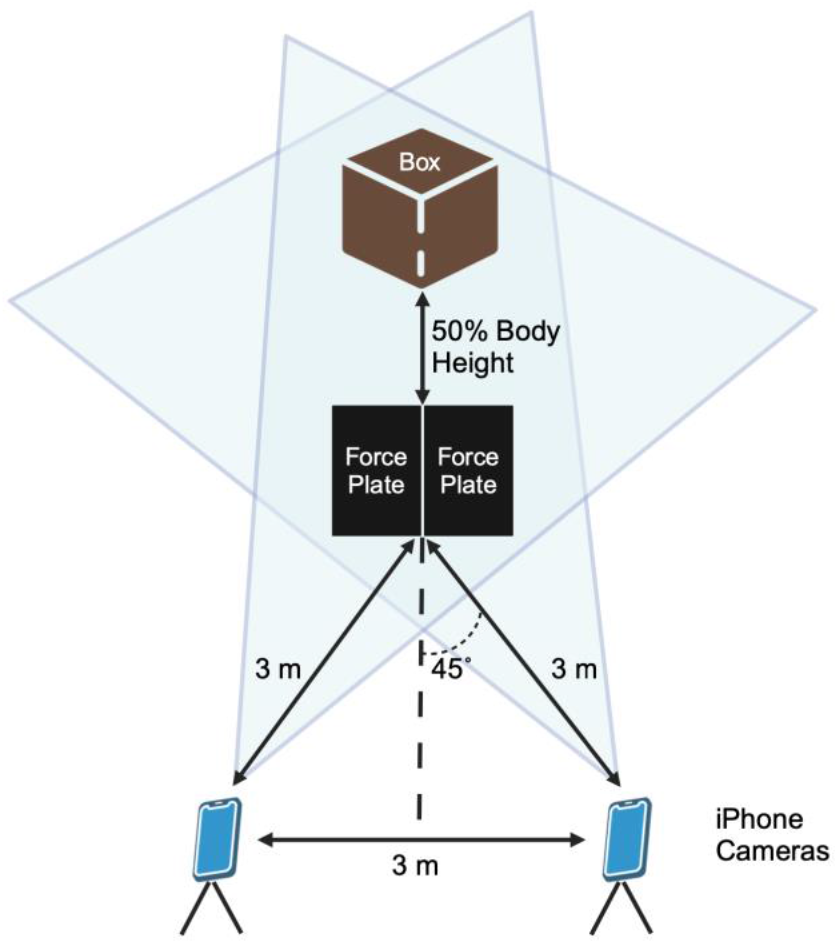
OpenCap Markerless Motion Capture and Force Plate Setup.

Vertical ground reaction forces (vGRF) were collected on a subsample of 12 participants within the post-ACLR cohort for validating event timing. The vGRF data was sampled at a frequency of 2,400 Hz from two embedded force plates (FP406020, Bertec Corp) and synchronized to the timing of the markerless motion capture system. The force plates were set up with a cartesian coordinate system with axes defined as *z*-axis vertical, *x*-axis anterior-posterior, and *y*-axis medial-lateral. Before starting each participant’s jump-landing trials, the force plates were zeroed, and the participant’s mass was recorded while they stood equally on each force plate.

#### The Pipeline: OpenLESS Automated Scoring

After recording jump-landing trials, raw trial data were automatically uploaded and processed over the cloud via OpenCap’s web interface. OpenCap’s automated processing suite included data extraction, pose estimation, time synchronization, and 3D anatomical marker set derivation.^7^ The 3D kinematics are then computed from the derived marker trajectories using inverse kinematics^25,26^ and a musculoskeletal model with biomechanical constraints.^7,27,28^ The resulting musculoskeletal model included 33 degrees of freedom (DOF) with 15 DOF used in our analysis – 3 DOF for the trunk, 3 DOF per hip, 1 DOF per knee, and 2 DOF per ankle.^7,27,28^ The final outputs of interest from the automated cloud processing included the marker (.trc) files containing 3D anatomical marker trajectories and motion (.mot) files containing 3D joint angles.

A custom Python (v.3.10.12) pipeline, OpenLESS, was developed for reading, signal processing, and extracting kinematic variables of interest from OpenCap, then algorithmically scoring the LESS based on jump-landing movement quality (Figure 4). OpenLESS Python script is provided in the Supplemental File 1. Before entry into the OpenLESS pipeline, each trial OpenCap video recording was inspected for completeness of the entirety of movement, and kinematic waveforms were assessed for biological plausibility. Trials were removed if there were any errors in cloud processing, pose estimation, and/or if the trial exuded excessive noise outside of expectation.^7,18,24^

**Figure 4.**
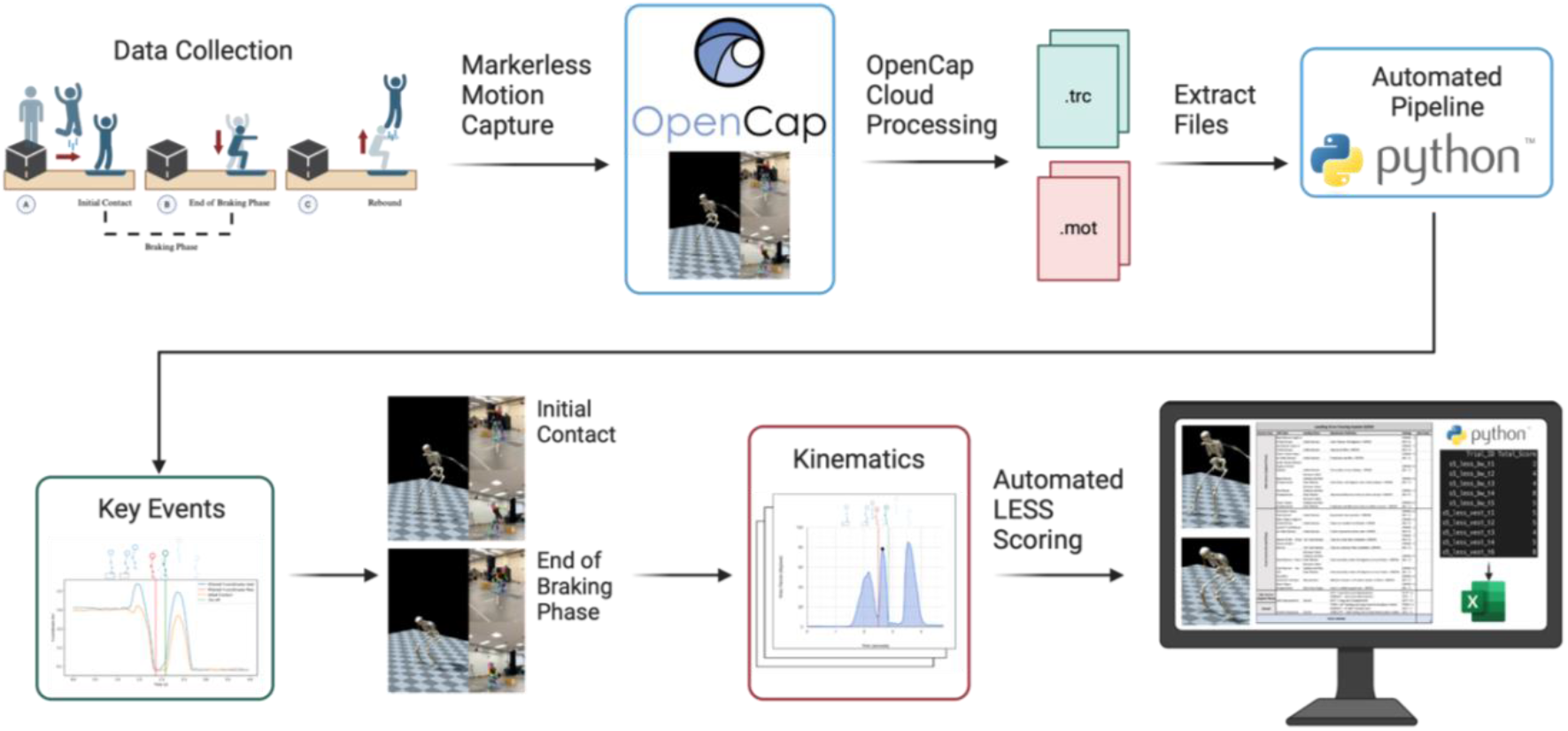
OpenLESS Automated Landing Error Scoring System Pipeline.

After determining eligibility, the marker trajectory and kinematic data were entered into the custom Python pipeline which performed the following. 1) marker trajectories and kinematics were filtered with a 4^th^ order, 12 Hz low-pass Butterworth filter; 2) stance phase was identified by determining key events for initial ground contact and rebound jump take-off; 3) first initial ground contact frame was identified as the first global minimum of the great toe marker trajectory in the vertical axis (*y*-axis); 4) rebound jump take-off frame was identified as the second global minimum of the great toe marker trajectory in the vertical axis; 5) a second quality control step was employed where the key events outlining the contact phase across the entire great toe marker trajectory in the vertical axis were plotted and inspected for accuracy; 6) the lowest point of the braking phase before upward movement was determined by the frame where knee flexion angle was at its peak; and 7) three-dimensional marker trajectories and joint angles at the initial contact and at the lowest point of the braking phase before upward movement frames were extracted.

The OpenLESS then uses the joint trajectories and kinematics at the event times of interest (Figure 2A to 2B) to score each item of the expanded 19-item LESS (22 possible errors) based on clinically- and literature-informed cut-offs.^3,5,9,12,13,21,29,30^ Identical to the expert rater LESS, an error present on either limb results in an error for that associated LESS item. The two additional OpenLESS items capture asymmetric foot landing and weight shift patterns, recently identified as clinically relevant movement characteristics.^13,30^ Technical details for scoring each LESS item are made available in Supplemental File 2. Data cleaning, processing, and LESS scoring were performed in Python (v3.10.12) using the *S*ci*P*y (v1.5.4), *pandas* (v2.2.2), and *NumPy* (v2.0) packages.

### Statistical Analysis

Statistical analyses were conducted using R Statistical Software (v4.4.1, R Core Team 2024). The normality of LESS scores was evaluated through visual inspection of histograms and quantile-quantile plots. Descriptive statistics are presented as mean ± standard deviation or median [interquartile range, IQR] if not normally distributed. Expert-rater LESS and automated OpenLESS scores were calculated as the mean score across all successful jump-landing trials within each participant.

Intraclass correlation coefficients (ICC) with 95% confidence intervals (CI) were calculated using the *psych* package (v2.6.4.26) to evaluate concurrent validity and reliability.^31,32^ A two-way random-effects model for an absolute agreement based on single ratings (ICC_2,1_) was used for event timing, and average ratings (ICC_2,k_) were used for LESS scores. For reliability, a linear mixed-effects model was used to calculate ICC_2,k_, optimizing the use of available information while accounting for variability across time points. ICC values were interpreted according to established guidelines: poor (0.0-0.5), moderate (0.5-0.75), good (0.75-0.9), and excellent (0.9-1.0).^32^

To further assess the validity of OpenLESS derived scores compared to expert-rater LESS scores, Pearson’s correlation coefficient (R) and Bland-Altman limits of agreement (LoA) were computed using the *stats* (v4.2.3) and *blandr* (v0.5.1) packages.^31^ Pearson correlations were categorized as per Portney and Watkins:^32^ ≤0.25 (little/no), 0.25-0.50 (low/fair), 0.50-0.75 (moderate/good), and ≥0.75 (strong) relationship.

Reliability measurement error was quantified using the standard error of measurement (SEM), calculated as 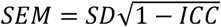, where SD represents the pooled standard deviation of test and retest scores.^31^ The minimal detectable change (MDC) was calculated as 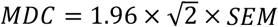.^31^ The SEM and MDC were expressed in the units of the measure (number of LESS errors).

## RESULTS

The healthy cohort used in the validation arm consisted of 26 individuals (12 males, 14 females; age = 23.0 ± 3.8 years, height = 171.9 ± 8.3 cm; mass = 75.4 ± 18.9 kg). The post-ACLR cohort used in the validation arm consisted of 27 individuals (8 males, 19 females; age = 21.4 ± 5.7 years, height = 173.5 ± 12.5 cm; mass = 73.9 ± 13.1 kg) that were 6-72 months post ACLR surgery (median: 33.0 [IQR: 50.5] months post-op; International Knee Documentation Committee [IKDC] = 83.2 ± 14.3). The field-based athlete cohort used in the reliability arm consisted of 39 recreationally athletic females (18 amateur soccer players, 10 university athletes from ball and non-ball sports, 11 recreational weightlifters; age = 25.0 ± 4.7 years, height = 165.0 ± 7.1 cm; mass = 63.5 ± 8.6 kg).

### Event Detection

The OpenLESS event detection pipeline, utilizing OpenCap kinematics and trajectories, demonstrated excellent validity in identifying initial ground contact and toe-off events when compared to force-plate measurements across 98 jump-landing trials (ICC_2,1_ > 0.99, p < 0.001, Supplemental File 3).

### Criterion Validity

Analysis of the healthy, college-aged cohort revealed good agreement between expert rater assessment and the automated OpenLESS pipeline for total LESS scores (ICC_2,*k*_ = 0.79, *p* < 0.001, Table 1). Box-whisker plots are presented in Figure 5A to display the range of values from both LESS scoring methods (Expert and OpenLESS). The Bland-Altman analysis (Figure 5B) estimated a mean bias of 0.35 (95% CI: -0.05, 0.75) indicating a small but non-significant systematic difference between the two methods (*t* = 1.80, *p* = 0.08). The LoA ranged from -1.59 (95% CI: -2.29, -0.90) to 2.30 (95% CI: 1.60, 2.99), representing that 95% of the differences between measurements fell within this range fall.

**Table 1.** Criterion Validity of OpenLESS in Healthy and Post-ACLR Cohorts.

| Cohort | Rater | Participants | Mean (SD) | ICC<sub>2,k</sub> |  |
| --- | --- | --- | --- | --- | --- |
|  |  |  |  | Value (95% CI) | R |
| Healthy Validation Cohort | OpenLESS | 26 | 2.78 (1.38) | 0.79 (0.55, 0.91)<sup>†</sup> | 0.70<sup>†</sup> |
|  | Expert |  | 3.13 (1.09) |  |  |
| Post-ACLR Validation Cohort | OpenLESS | 27 | 5.50 (3.47) | 0.88 (0.55, 0.96)<sup>†</sup> | 0.86<sup>†</sup> |
|  | Expert |  | 7.01 (3.71) |  |  |

Notes: Averaged composite LESS scores computed from automated OpenLESS compared against an expert rater in a healthy (12 males and 14 females) and post-ACLR (8 males and 19 females) sample. Criterion validity was calculated using a linear mixed-effects model with the ICC function in the *psych* package (v2.6.4.26).
Confidence interval, CI; intraclass correlation coefficient, ICC; Landing Error Scoring System, LESS.
Pearson's correlation coefficient, R.
<sup>†</sup> Statistically significant *p*-value <.05.

**Figure 5.**
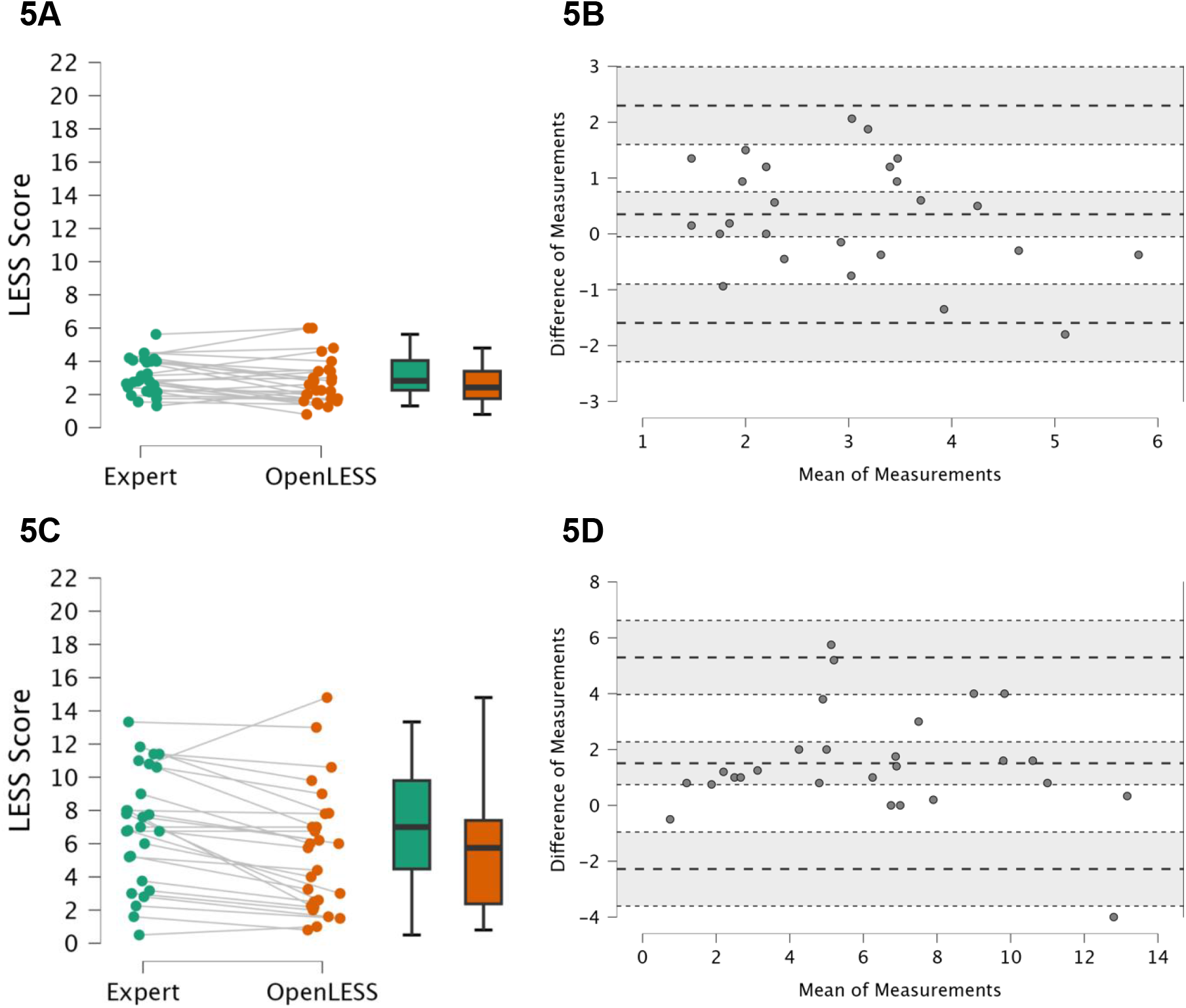
(A) Healthy cohort box and whisker plot of LESS scores by both methods. (B) Healthy cohort Bland-Altman plot of OpenLESS compared to expert rater. (C) Post-ACLR cohort box and whisker plot of LESS scores by both methods. (D) Post-ACLR cohort Bland-Altman plot of OpenLESS compared to expert rater. The x-axis “Means” refers to the mean score from the two rating systems (OpenLESS and expert). The y-axis “Differences” refers to the difference in score between the two rating systems, i.e., expert rater minus OpenLESS. Mean bias is represented by the central dashed line, whereas the upper and lower dashed lines represent the upper and lower limits of agreement. 95% confidence intervals are indicated by the shaded regions surrounding mean bias and the limits of agreement.

Good agreement was observed in the clinically relevant post-ACLR cohort (ICC_2,*k*_ = 0.88, *p* < 0.001, Table 1). Box-whisker plots are presented in Figure 5C to display the range of values from both LESS scoring methods (Expert and OpenLESS). In the Bland-Altman analysis (Figure 5D), a significant (*t* = 4.06, *p* < 0.001), directionally similar systematic bias was observed in the post-ACLR cohort compared to the healthy cohort, where OpenLESS consistently scored lower than the expert rater with a mean bias of 1.51 errors (95% CI: 0.74, 2.27). The LoA ranged from -2.28 (95% CI: -3.60, -0.96) to 5.30 (95% CI: 3.97, 6.62).

### Intersession Reliability

A repeated measures assessment of the OpenLESS score was conducted across four visits, with 17 participants attending all four visits, 19 attending three, and 3 attending two visits. Mean OpenLESS scores demonstrated minimal variation, ranging from 5.35 errors at visit one, 5.12 at visit two, 5.87 at visit three, and 5.00 at visit four (Figure 6).

**Figure 6.**
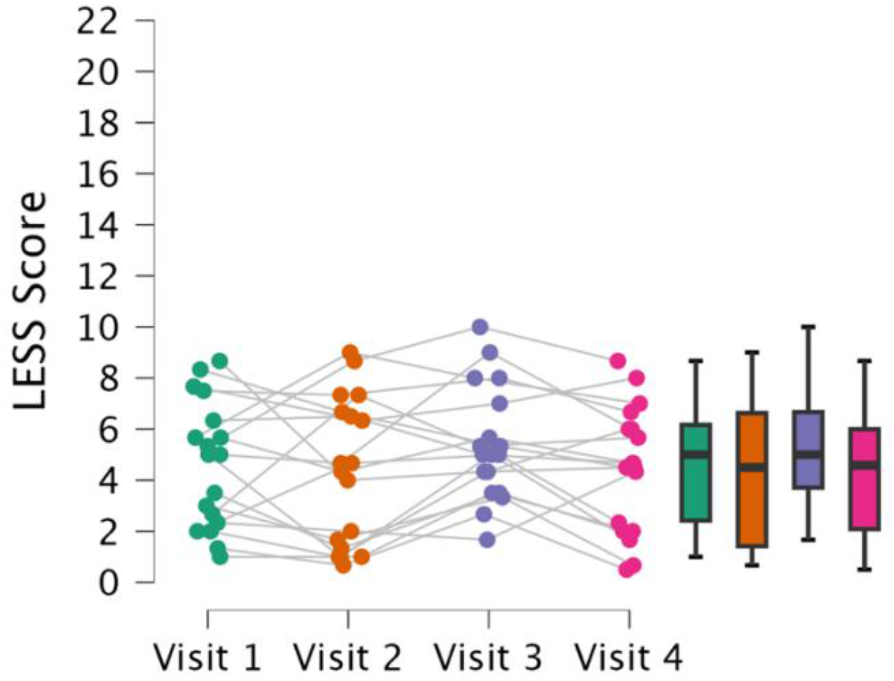
Field-Based Athlete Cohort LESS scores graded by OpenLESS. Box and whisker plots with jittered athlete LESS scores across the four repeated visits.

OpenLESS demonstrated good to excellent intersession reliability (ICC_2,*k*_ = 0.89, p < 0.001, Table 2). Measurement error metrics indicated an SEM of 0.98 and an MDC of 2.72 OpenLESS errors.

**Table 2.** Intersession Reliability of OpenLESS in Field-Based Athlete Cohort.

| Cohort | Session | Participants | Mean (SD) | ICC<sub>2,k</sub> |  |  |
| --- | --- | --- | --- | --- | --- | --- |
|  |  |  |  | Value (95% CI) | SEM | MDC |
| Athlete<br>Reliability<br>Cohort | Visit 1 | 39 | 5.35 (2.84) | 0.89 (0.81, 0.93)<sup>†</sup> | 0.98 | 2.72 |
|  | Visit 2 | 35 | 5.12 (3.20) |  |  |  |
|  | Visit 3 | 34 | 5.87 (2.44) |  |  |  |
|  | Visit 4 | 23 | 5.00 (2.65) |  |  |  |

*Notes:* Averaged composite LESS scores computed from automated OpenLESS from up to 4 visits (each 7 days apart) in 39 female athletes, all athletes contributed at least two visits. Intersession reliability was calculated using a linear mixed-effects model with the ICC function in the *psych* package (v2.6.4.26).
Confidence interval, CI; intraclass correlation coefficient, ICC; Landing Error Scoring System, LESS; minimal detectable change, MDC; standard error of measurement, SEM.
<sup>†</sup> Statistically significant *p*-value <.05.

## DISCUSSION

This study developed and validated OpenLESS (Supplemental File 1), an automated scoring system for the LESS that leverages markerless motion capture technology to enhance the efficiency and scalability of movement quality assessment. OpenLESS demonstrated good agreement with expert raters across healthy and post-ACLR cohorts while showing excellent intersession reliability in a field-based athletic cohort. The kinematic-based event detection algorithm for identifying the landing phase also showed excellent validity against gold-standard force plate measurements. Building upon previous efforts to automate movement quality assessments,^2,3^ OpenLESS provides clinicians and researchers with an efficient, accessible tool for capturing and grading jump-landing mechanics.

A fundamental validation step for the OpenLESS pipeline was establishing the accuracy of its event detection algorithm, which processes markerless motion data from OpenCap to identify key jump-landing events (initial contact and toe-off). We compared OpenLESS’s automated event detection against gold-standard force plate measurements in 98 trials from 12 post-ACLR participants. The analysis revealed near-perfect agreement (ICC_2,1_ > 0.99) between OpenLESS and force plate event timing, validating our approach to automated event detection using markerless motion capture data. This high level of agreement demonstrates that OpenLESS can reliably identify key biomechanical events without requiring specialized laboratory equipment.

OpenLESS demonstrated good agreement with expert raters in both healthy and post-ACLR cohorts, though with a consistent, systematic bias toward slightly lower scores, particularly in the post-ACLR group. This bias likely reflects differences between algorithmic and human movement quality assessment thresholds, a pattern commonly observed in automated kinematic analysis systems.^33^ Notably, the agreement between OpenLESS and expert raters (ICC_2,*k*_ = 0.79-0.88) closely parallels the documented interrater reliability among expert raters themselves (ICC_2,*k*_ = 0.83-0.92), suggesting that OpenLESS’s scoring variability falls within acceptable clinical limits.^9,10,34^ This reliability is particularly significant given that small sample sizes often constrain biomechanical studies investigating injury risk and surgical outcomes due to resource limitations.^35^ The automated nature of OpenLESS could help address these constraints by enabling larger-scale assessments while maintaining measurement consistency.

OpenLESS demonstrated robust scoring consistency across multiple testing environments, including an outdoor soccer pitch, athletic field, and indoor recreation center, validating its utility for movement quality assessment beyond traditional laboratory settings. Score stability was evident across visits with acceptable measurement error.^10,31^ This temporal stability is essential for monitoring interventions and rehabilitation progress, where reliable baseline measurements enable detection of meaningful change.^36^ While traditional LESS scoring can be subject to rater variability,^10^ OpenLESS achieves high intersession reliability (ICC_2,*k*_ = 0.89), enhancing precision for longitudinal and post-intervention assessments. This precision aligns with the growing emphasis on evidence-based injury prevention protocols.^1–4^ The minimal measurement error further establishes OpenLESS for use in clinical trials and rehabilitation, where repeated assessments are essential for evaluating improvements and the effectiveness of intervention programs.

The assessment of OpenLESS’s measurement properties across three distinct participant populations demonstrates its robust clinical utility and versatility in real-world settings, directly addressing critical needs in lower extremity injury risk prevention and intervention.^1–4^ Unlike traditional research that prioritizes internal validity through strict inclusion/exclusion criteria,^36^ our approach deliberately embraced ecological validity by testing diverse populations and environments. We validated OpenLESS not only in healthy individuals but also in people post-ACLR, providing evidence of its effectiveness in clinically relevant populations. Our reliability testing further emphasized real-world applicability by conducting assessments outside laboratory environments and allowing participants to wear their typical exercise footwear and clothing—conditions known to challenge markerless motion capture systems but essential for clinical implementation.^6,37^

OpenCap, the foundation of our system, has proven its versatility across multiple applications, from analyzing gait in neurological disorders^38^ to assessing lower extremity kinematics during cycling^39^ and evaluating whole-body movement during dynamic balance tasks.^40^ By building OpenLESS as an open-source tool on this established platform, we provide researchers and clinicians with an adaptable framework that can be customized for various dynamic activities and populations, while maintaining measurement precision in real-world conditions. This accessibility directly responds to the documented need for reliable movement assessment tools that can be deployed beyond laboratory settings.^13,37^ The combination of OpenCap and OpenLESS creates new opportunities for larger-scale studies and widespread clinical implementation of sophisticated biomechanical analysis,^7,37,41^ potentially transforming how we approach movement screening and injury prevention across diverse populations.

While previous approaches have employed machine learning to predict LESS total scores based on expert grader patterns,^15^ OpenLESS takes a fundamentally different approach. Rather than replicating human scoring patterns, OpenLESS directly processes motion capture data to evaluate LESS items using categorical criteria, aligning with the original LESS development methodology.^5,9^ This transparent approach, combined with accessible source code, addresses the implementation limitations often encountered with proprietary assessment tools.

Biomechanical injury risk assessments have faced criticism for their narrow focus.^42^ Injury risk, both primary and secondary, involves multiple factors beyond physical function and performance metrics.^43,44^ Therefore, OpenLESS should be integrated within comprehensive biopsychosocial assessments for complete performance characterization.^45^ While OpenLESS effectively identifies aberrant movement patterns consistent with the original LESS,^5,9^ such as decreased hip and knee flexion during landing, several limitations warrant consideration. OpenLESS demonstrated a small systematic bias toward lower scores than expert raters, suggesting more stringent grading criteria for aberrant movement patterns. The MDC across sessions was slightly larger for OpenLESS (2.72 errors) compared to expert raters (2.25 errors),^10^ though this comparison should be interpreted cautiously given the different sample sizes (n = 39 versus n = 13, respectively). Additionally, while our validity cohorts included both male and female participants, the reliability analyses were conducted exclusively in female athletes, limiting the generalizability of the reliability findings across the sexes.

Future development should continue leveraging open-source tools like OpenCap to enhance accessibility and automate additional clinical and return-to-activity assessments, including cutting movement assessment scores and single-leg LESS variations. Priority areas for future research include validating OpenLESS in other clinically relevant populations and expanding validation studies to larger healthy cohorts, with a particular focus on intervention responsiveness. Encouraging open-source tools not only increases accessibility but also empowers clinicians and researchers of all experience levels to utilize and adapt these methods for diverse clinical and research applications.

## CONCLUSION

OpenLESS provides a comprehensive, automated pipeline for jump-landing assessment, encompassing data processing, event detection, and standardized LESS scoring based on established operational definitions. The system demonstrates robust reliability and clinical utility across diverse settings, offering time-efficient movement quality assessment without specialized laboratory equipment. Initial validation in healthy college-aged and post-ACLR populations supports OpenLESS as a promising tool for democratizing evidence-based injury risk screening. By reducing barriers to implementation while maintaining measurement precision, OpenLESS advances the field toward more accessible, standardized biomechanical assessment.

## Data Availability

All data produced in the present study are available upon reasonable request to the authors.

## Acknowledgments

The authors thank Adrian Pang, Sam Noel, Siam Kirby, and Skyler Peterson, who assisted in the data collection of this study. Additionally, the authors thank the participants who agreed to volunteer for this study. The views and opinions expressed are those of the authors and do not reflect the official policy of the Department of the Army, the Department of Defense, or the United States Government.

## REFERENCES

1. Hewett TE, Bates NA. Preventive Biomechanics: A Paradigm Shift With a Translational Approach to Injury Prevention. Am J Sports Med. 2017;45(11):2654–2664. doi:10.1177/0363546516686080

2. Padua DA, DiStefano LJ, Hewett TE, et al. National Athletic Trainers’ Association Position Statement: Prevention of Anterior Cruciate Ligament Injury. J Athl Train. 2018;53(1):5–19. doi:10.4085/1062-6050-99

3. Whittaker JL, Booysen N, de la Motte S, et al. Predicting sport and occupational lower extremity injury risk through movement quality screening: a systematic review. Br J Sports Med. 2017;51(7):580–585. doi:10.1136/bjsports-2016-096760

4. Logerstedt DS, Scalzitti D, Risberg MA, et al. Knee Stability and Movement Coordination Impairments: Knee Ligament Sprain Revision 2017. J Orthop Sports Phys Ther. 2017;47(11):A1–A47. doi:10.2519/jospt.2017.0303

5. Padua DA, DiStefano LJ, Beutler AI, de la Motte SJ, DiStefano MJ, Marshall SW. The Landing Error Scoring System as a Screening Tool for an Anterior Cruciate Ligament Injury-Prevention Program in Elite-Youth Soccer Athletes. J Athl Train. 2015;50(6):589–595. doi:10.4085/1062-6050-50.1.10

6. Colyer SL, Evans M, Cosker DP, Salo AIT. A Review of the Evolution of Vision-Based Motion Analysis and the Integration of Advanced Computer Vision Methods Towards Developing a Markerless System. Sports Med - Open. 2018;4(1):24. doi:10.1186/s40798-018-0139-y

7. Uhlrich SD, Falisse A, Kidziński Ł, et al. OpenCap: Human movement dynamics from smartphone videos. PLOS Comput Biol. 2023;19(10):e1011462. doi:10.1371/journal.pcbi.1011462

8. Lally EM, Ericksen H, Earl-Boehm J. Measurement Properties of Clinically Accessible Movement Assessment Tools for Analyzing Jump Landings: A Systematic Review. J Sport Rehabil. 2022;31(4):465–475. doi:10.1123/jsr.2021-0288

9. Padua DA, Marshall SW, Boling MC, Thigpen CA, Garrett WE, Beutler AI. The Landing Error Scoring System (LESS) Is a valid and reliable clinical assessment tool of jump-landing biomechanics: The JUMP-ACL study. Am J Sports Med. 2009;37(10):1996–2002. doi:10.1177/0363546509343200

10. Hanzlíková I, Hébert-Losier K. Is the Landing Error Scoring System Reliable and Valid? A Systematic Review. Sports Health. 2020;12(2):181–188. doi:10.1177/1941738119886593

11. Onate J, Cortes N, Welch C, Van Lunen BL. Expert versus novice interrater reliability and criterion validity of the landing error scoring system. J Sport Rehabil. 2010;19(1):41–56. doi:10.1123/jsr.19.1.41

12. Padua DA, Distefano LJ. Sagittal Plane Knee Biomechanics and Vertical Ground Reaction Forces Are Modified Following ACL Injury Prevention Programs: A Systematic Review. Sports Health. 2009;1(2):165–173. doi:10.1177/1941738108330971

13. Mauntel TC, Padua DA, Stanley LE, et al. Automated Quantification of the Landing Error Scoring System With a Markerless Motion-Capture System. J Athl Train. 2017;52(11):1002–1009. doi:10.4085/1062-6050-52.10.12

14. Mauntel TC, Cameron KL, Pietrosimone B, Marshall SW, Hackney AC, Padua DA. Validation of a Commercially Available Markerless Motion-Capture System for Trunk and Lower Extremity Kinematics During a Jump-Landing Assessment. J Athl Train. 2021;56(2):177–190. doi:10.4085/1062-6050-0023.20

15. Hébert-Losier K, Hanzlíková I, Zheng C, Streeter L, Mayo M. The ‘DEEP’ Landing Error Scoring System. Appl Sci. 2020;10(3):892. doi:10.3390/app10030892

16. Cao Z, Hidalgo G, Simon T, Wei SE, Sheikh Y. OpenPose: Realtime Multi-Person 2D Pose Estimation Using Part Affinity Fields. IEEE Trans Pattern Anal Mach Intell. 2021;43(1):172–186. doi:10.1109/TPAMI.2019.2929257

17. Sun K, Xiao B, Liu D, Wang J. Deep High-Resolution Representation Learning for Human Pose Estimation. Published online February 25, 2019. doi:10.48550/arXiv.1902.09212

18. Turner JA, Chaaban CR, Padua DA. Validation of OpenCap: A low-cost markerless motion capture system for lower-extremity kinematics during return-to-sport tasks. J Biomech. 2024;171:112200. doi:10.1016/j.jbiomech.2024.112200

19. Vandenbroucke JP, von Elm E, Altman DG, et al. Strengthening the Observational Studies Reporting of in Epidemiology (STROBE): explanation and elaboration. PLoS Med. 2007;4(10):e297. doi:10.1371/journal.pmed.0040297

20. Horsak B, Prock K, Krondorfer P, Siragy T, Simonlehner M, Dumphart B. Inter-trial variability is higher in 3D markerless compared to marker-based motion capture: Implications for data post-processing and analysis. J Biomech. 2024;166:112049. doi:10.1016/j.jbiomech.2024.112049

21. Turner JA, Hartshorne M, Cameron KL, Padua D. Jump Landing Kinematics: Establishing Normative Ranges For Male And Female Athletes. J Athl Train. Published online April 18, 2024. doi:10.4085/1062-6050-0006.24

22. Peng HT. Changes in Biomechanical Properties during Drop Jumps of Incremental Height. J Strength Cond Res. 2011;25(9):2510. doi:10.1519/JSC.0b013e318201bcb3

23. OpenCap - Musculoskeletal forces from smartphone videos. OpenCap. 2022. Accessed July 6, 2024. https://www.opencap.ai/

24. Horsak B, Eichmann A, Lauer K, et al. Concurrent validity of smartphone-based markerless motion capturing to quantify lower-limb joint kinematics in healthy and pathological gait. J Biomech. 2023;159:111801. doi:10.1016/j.jbiomech.2023.111801

25. Delp SL, Anderson FC, Arnold AS, et al. OpenSim: Open-Source Software to Create and Analyze Dynamic Simulations of Movement. IEEE Trans Biomed Eng. 2007;54(11):1940–1950. doi:10.1109/TBME.2007.901024

26. Seth A, Hicks JL, Uchida TK, et al. OpenSim: Simulating musculoskeletal dynamics and neuromuscular control to study human and animal movement. PLOS Comput Biol. 2018;14(7):e1006223. doi:10.1371/journal.pcbi.1006223

27. Lai AKM, Arnold AS, Wakeling JM. Why are Antagonist Muscles Co-activated in My Simulation? A Musculoskeletal Model for Analysing Human Locomotor Tasks. Ann Biomed Eng. 2017;45(12):2762–2774. doi:10.1007/s10439-017-1920-7

28. Rajagopal A, Dembia CL, DeMers MS, Delp DD, Hicks JL, Delp SL. Full-Body Musculoskeletal Model for Muscle-Driven Simulation of Human Gait. IEEE Trans Biomed Eng. 2016;63(10):2068–2079. doi:10.1109/TBME.2016.2586891

29. Bishop C, Jordan M, Torres-Ronda L, et al. Selecting Metrics That Matter: Comparing the Use of the Countermovement Jump for Performance Profiling, Neuromuscular Fatigue Monitoring, and Injury Rehabilitation Testing. Strength Cond J. 2023;45(5):545. doi:10.1519/SSC.0000000000000772

30. Eckard TG, Miraldi SFP, Peck KY, et al. Automated Landing Error Scoring System Performance and the Risk of Bone Stress Injury in Military Trainees. J Athl Train. 2021;57(4):334–340. doi:10.4085/1062-6050-0263.21

31. de Vet HCW, Terwee CB, Mokkink LB, Knol DL. Measurement in Medicine: A Practical Guide. Cambridge University Press; 2011. doi:10.1017/CBO9780511996214

32. Portney LG, Watkins MP. Foundations of Clinical Research: Applications to Practice. Pearson/Prentice Hall; 2009.

33. Breithaupt K, Hare D. Automated Test Assembly. In: Technology and Testing. Routledge; 2015.

34. Padua DA, Boling MC, Distefano LJ, Onate JA, Beutler AI, Marshall SW. Reliability of the landing error scoring system-real time, a clinical assessment tool of jump-landing biomechanics. J Sport Rehabil. 2011;20(2):145–156. doi:10.1123/jsr.20.2.145

35. Knudson DV. Authorship and sampling practice in selected biomechanics and sports science journals. Percept Mot Skills. 2011;112(3):838–844. doi:10.2466/17.PMS.112.3.838-844

36. Ho PM, Peterson PN, Masoudi FA. Evaluating the Evidence. Circulation. 2008;118(16):1675–1684. doi:10.1161/CIRCULATIONAHA.107.721357

37. Armitano-Lago C, Willoughby D, Kiefer AW. A SWOT Analysis of Portable and Low-Cost Markerless Motion Capture Systems to Assess Lower-Limb Musculoskeletal Kinematics in Sport. Front Sports Act Living. 2022;3:809898. doi:10.3389/fspor.2021.809898

38. Min YS, Jung TD, Lee YS, et al. Biomechanical Gait Analysis Using a Smartphone-Based Motion Capture System (OpenCap) in Patients with Neurological Disorders. Bioengineering. 2024;11(9):911. doi:10.3390/bioengineering11090911

39. Kakavand R, Ahmadi R, Parsaei A, Edwards WB, Komeili A. OpenCap markerless motion capture estimation of lower extremity kinematics and dynamics in cycling. Published online August 20, 2024. doi:10.48550/arXiv.2409.03766

40. Turner JA, Hartshorne ML, Padua DA. Role of Thigh Muscle Strength and Joint Kinematics in Dynamic Stability: Implications for Y-Balance Test Performance. J Sport Rehabil. 2024;33(8):654–662. doi:10.1123/jsr.2024-0081

41. Heron M, Hanson VL, Ricketts I. Open source and accessibility: advantages and limitations. J Interact Sci. 2013;1(1):2. doi:10.1186/2194-0827-1-2

42. Bittencourt NFN, Meeuwisse WH, Mendonça LD, Nettel-Aguirre A, Ocarino JM, Fonseca ST. Complex systems approach for sports injuries: moving from risk factor identification to injury pattern recognition—narrative review and new concept. Br J Sports Med. 2016;50(21):1309–1314. doi:10.1136/bjsports-2015-095850

43. Andersen MB, Williams JM. A Model of Stress and Athletic Injury: Prediction and Prevention. J Sport Exerc Psychol. 1988;10(3):294–306. doi:10.1123/jsep.10.3.294

44. Van Eetvelde H, Mendonça LD, Ley C, Seil R, Tischer T. Machine learning methods in sport injury prediction and prevention: a systematic review. J Exp Orthop. 2021;8(1):27. doi:10.1186/s40634-021-00346-x

45. McClean ZJ, Pasanen K, Lun V, et al. A Biopsychosocial Model for Understanding Training Load, Fatigue, and Musculoskeletal Sport Injury in University Athletes: A Scoping Review. J Strength Cond Res. 2024;38(6):1177. doi:10.1519/JSC.0000000000004789

