## Supplemental File 1 for "Open Source, Open Science: Development of OpenLESS as the Automated Landing Error Scoring System"

### %% [markdown]
### # OpenCap Pipeline for Automating Landing Error Scoring System
#### START AT Line
### %% [markdown]
### This pipeline will read in a number of .csv and outputs a new dataframe with rows as observations and columns as scores
#
### %% [markdown]
### ## Preamable
# %%
### -\*- coding: utf-8 -\*-
import numpy as np
import matplotlib.pyplot as plt
import pandas as pd
import os
from prettytable import PrettyTable
from scipy import signal
from scipy.signal import butter, filtfilt, argrelmin
column\_headers = [
"Trial\_ID",
"Total\_Score",
"1\_Knee\_Flexion\_IC\_L",
"1\_Knee\_Flexion\_IC\_R",
"1\_Knee\_Flexion\_IC\_T",
"2\_Hip\_Flexion\_IC\_L",
"2\_Hip\_Flexion\_IC\_R",
"2\_Hip\_Flexion\_IC\_T",
"3\_Trunk\_Flexion\_IC\_T",
"4\_Ankle\_Plantar\_Flexion\_IC\_L",
"4\_Ankle\_Plantar\_Flexion\_IC\_R",
"4\_Ankle\_Plantar\_Flexion\_IC\_T",
"5\_Asymmetrical\_Timing",
"6\_Asymmetrical\_HeelToe\_ToeHeel",
"7\_Lateral\_Trunk\_Flexion\_IC\_L",
"7\_Lateral\_Trunk\_Flexion\_IC\_R",
"7\_Lateral\_Trunk\_Flexion\_IC\_T",
"8\_Medial\_Knee\_IC\_L",
"8\_Medial\_Knee\_IC\_R",
"8\_Medial\_Knee\_IC\_T",
"9\_Stance\_Wide\_IC\_L",
"9\_Stance\_Wide\_IC\_R",
"9\_Stance\_Wide\_IC\_T",
"10\_Stance\_Narrow\_IC\_L",
"10\_Stance\_Narrow\_IC\_R",
"10\_Stance\_Narrow\_IC\_T",
"11\_IR\_Foot\_IC\_L",
"11\_IR\_Foot\_IC\_R",
"11\_IR\_Foot\_IC\_T",
"12\_ER\_Foot\_IC\_L",
"12\_ER\_Foot\_IC\_R",
"12\_ER\_Foot\_IC\_T",
"13\_Knee\_Flexion\_Displacement\_L",
"13\_Knee\_Flexion\_Displacement\_R",
"13\_Knee\_Flexion\_Displacement\_T",
"14\_Hip\_Flexion\_Displacement\_L",
"14\_Hip\_Flexion\_Displacement\_R",
"14\_Hip\_Flexion\_Displacement\_T",
"15\_Trunk\_Flexion\_Max\_T",
"16\_Medial\_Knee\_Max\_L",
"16\_Medial\_Knee\_Max\_R",
"16\_Medial\_Knee\_Max\_T",
"17\_Asymmetrical\_Load\_T",
"18\_Joint\_Displacement\_T",
"19\_Overall\_Impression\_T",
"1\_Knee\_Flexion\_IC\_L\_deg",
"1\_Knee\_Flexion\_IC\_R\_deg",
"2\_Hip\_Flexion\_IC\_L\_deg",
"2\_Hip\_Flexion\_IC\_R\_deg",
"3\_Trunk\_Flexion\_IC\_deg",
"7\_Lateral\_Trunk\_Flexion\_IC\_deg",
"8\_Medial\_Knee\_IC\_L\_deg",
"8\_Medial\_Knee\_IC\_R\_deg",
"9\_Stance\_Hip\_abd\_IC\_L\_deg",
"9\_Stance\_Hip\_abd\_IC\_R\_deg",
"11\_IR\_Foot\_IC\_L\_deg",
"11\_IR\_Foot\_IC\_R\_deg",
"12\_ER\_Foot\_IC\_L\_deg",
"12\_ER\_Foot\_IC\_R\_deg",
"13\_Knee\_Flexion\_Displacement\_L\_deg",
"13\_Knee\_Flexion\_Displacement\_R\_deg",
"14\_Hip\_Flexion\_Displacement\_L\_deg",
"14\_Hip\_Flexion\_Displacement\_R\_deg",
"15\_Trunk\_Flexion\_Max\_deg",
"16\_Medial\_Knee\_Max\_L\_deg",
"16\_Medial\_Knee\_Max\_R\_deg",
"17\_Asymmetrical\_Load\_L\_deg",
"17\_Asymmetrical\_Load\_R\_deg",
]
LESS\_results = pd.DataFrame(columns=column\_headers)
print(LESS\_results)
column\_headers\_short = [
"Trial\_ID",
"Total\_Score",
"1\_Knee\_Flexion\_IC\_T",
"2\_Hip\_Flexion\_IC\_T",
"3\_Trunk\_Flexion\_IC\_T",
"4\_Ankle\_Plantar\_Flexion\_IC\_T",
"5\_Asymmetrical\_Timing",
"6\_Asymmetrical\_HeelToe\_ToeHeel",
"7\_Lateral\_Trunk\_Flexion\_IC\_T",
"8\_Medial\_Knee\_IC\_T",
"9\_Stance\_Wide\_IC\_T",
"10\_Stance\_Narrow\_IC\_T",
"11\_IR\_Foot\_IC\_T",
"12\_ER\_Foot\_IC\_T",
"13\_Knee\_Flexion\_Displacement\_T",
"14\_Hip\_Flexion\_Displacement\_T",
"15\_Trunk\_Flexion\_Max\_T",
"16\_Medial\_Knee\_Max\_T",
"17\_Asymmetrical\_Load\_T",
"18\_Joint\_Displacement\_T",
"19\_Overall\_Impression\_T",
]
LESS\_results\_short = pd.DataFrame(columns=column\_headers\_short)
for index, header in enumerate(column\_headers):
print(f"Column {index}: {header}")
# %%
def load\_coord\_file(file\_name):
data = np.loadtxt(file\_name, skiprows=6)
time = data[:, 1]
y\_coords\_left\_toes = data[:, 45]
y\_coords\_left\_heel = data[:, 51]
y\_coords\_right\_toes = data[:, 54]
y\_coords\_right\_heel = data[:, 60]
return (
data,
time,
y\_coords\_left\_toes,
y\_coords\_left\_heel,
y\_coords\_right\_toes,
y\_coords\_right\_heel,
)
def apply\_butterworth\_filter\_coords(
y\_coords\_left\_toes,
y\_coords\_left\_heel,
y\_coords\_right\_toes,
y\_coords\_right\_heel,
sampling\_freq,
coord\_cutoff\_freq,
):
cutoff\_freq = coord\_cutoff\_freq # Hz
fs = sampling\_freq # Sampling frequency
nyquist\_freq = fs / 2 # Nyquist frequency
filter\_order = 4 # Filter order
b, a = butter(filter\_order, cutoff\_freq / nyquist\_freq, btype="low")
left\_toes\_filtered = filtfilt(b, a, y\_coords\_left\_toes)
left\_heel\_filtered = filtfilt(b, a, y\_coords\_left\_heel)
right\_toes\_filtered = filtfilt(b, a, y\_coords\_right\_toes)
right\_heel\_filtered = filtfilt(b, a, y\_coords\_right\_heel)
return (
left\_toes\_filtered,
left\_heel\_filtered,
right\_toes\_filtered,
right\_heel\_filtered,
)
def apply\_butterworth\_filter(file\_path, sampling\_freq, cutoff\_freq):
if isinstance(file\_path, pd.DataFrame):
data = file\_path
else:
data = pd.read\_csv(file\_path)
filtered\_data = data.copy()
for column in data.columns:
if column != "time":
column\_data = data[column]
cutoff\_freq = cutoff\_freq # Hz
fs = sampling\_freq # Sampling frequency
nyquist\_freq = fs / 2 # Nyquist frequency
filter\_order = 4 # Filter order
b, a = butter(filter\_order, cutoff\_freq / nyquist\_freq, btype="low")
padlen = len(column\_data) // 2 # Adjust padlen according to your needs
filtered\_column\_data = filtfilt(b, a, column\_data, padlen=padlen)
filtered\_data[column] = filtered\_column\_data
return filtered\_data
def id\_initialcontact\_toes(toes\_filtered, time):
toes\_local\_minima\_indices = argrelmin(toes\_filtered)
start\_threshold = toes\_filtered[0] - 0.2
first\_minima\_index = None
for index in toes\_local\_minima\_indices[0]:
if toes\_filtered[index] < start\_threshold:
first\_minima\_index = index
break
toes\_first\_minima\_time = (
time[first\_minima\_index] if first\_minima\_index is not None else None
)
toes\_first\_minima\_y\_coord = (
toes\_filtered[first\_minima\_index] if first\_minima\_index is not None else None
)
return toes\_first\_minima\_time, toes\_first\_minima\_y\_coord, first\_minima\_index
def id\_initialcontact\_heels(heel\_filtered, time):
heel\_local\_minima\_indices = argrelmin(heel\_filtered)
start\_threshold = heel\_filtered[0] - 0.2
first\_minima\_index = None
for index in heel\_local\_minima\_indices[0]:
if heel\_filtered[index] < start\_threshold:
first\_minima\_index = index
break
heel\_first\_minima\_time = (
time[first\_minima\_index] if first\_minima\_index is not None else None
)
heel\_first\_minima\_y\_coord = (
heel\_filtered[first\_minima\_index] if first\_minima\_index is not None else None
)
return heel\_first\_minima\_time, first\_minima\_index
### %% [markdown]
### ### Scoring Function
# %%
def process\_bilateral\_LESS(
right\_trial\_file,
left\_trial\_file,
coord\_file,
LESS\_results,
sampling\_freq,
coord\_cutoff\_freq,
):
input\_df\_r = pd.read\_csv(right\_trial\_file)
input\_df\_l = pd.read\_csv(left\_trial\_file)
Trial\_ID = right\_trial\_file.replace("right\_kin\_", "").replace("\_filtered.csv", "")
LESS\_results.at[Trial\_ID, "Trial\_ID"] = Trial\_ID
LESS\_results\_short.at[Trial\_ID, "Trial\_ID"] = Trial\_ID
"""
----------------------------------------------------------------------------
KNEE FLEXION
1. INITIAL CONTACT
13. MAXIMUM POSITION
"""
(
max\_knee\_flexion\_deg\_r,
max\_knee\_flexion\_deg\_l,
ic\_knee\_flexion\_total,
max\_knee\_flexion\_total,
) = id\_knee\_flexion\_error(Trial\_ID, input\_df\_r, input\_df\_l)
"""
----------------------------------------------------------------------------
HIP FLEXION
----------------------------------------------------------------------------
2. INITIAL CONTACT
14. MAXIMUM POSITION
"""
(
max\_hip\_flexion\_deg\_r,
max\_hip\_flexion\_deg\_l,
ic\_hip\_flexion\_total,
max\_hip\_flexion\_total,
) = id\_hip\_flexion\_error(Trial\_ID, input\_df\_r, input\_df\_l)
"""
----------------------------------------------------------------------------
TRUNK FLEXION
----------------------------------------------------------------------------
3. INITIAL CONTACT
15. MAXIMUM POSITION
"""
ic\_trunk\_flexion\_total, max\_trunk\_flexion\_total = id\_trunk\_flexion\_error(
Trial\_ID, input\_df\_r
)
"""
----------------------------------------------------------------------------
ANKLE PLANTARFLEXION
----------------------------------------------------------------------------
4. INITIAL CONTACT
"""
ankle\_PF\_left, ankle\_PF\_right, ic\_ankle\_PF\_total = id\_ankle\_plantarflexion\_error(
Trial\_ID, coord\_file, sampling\_freq, coord\_cutoff\_freq
)
"""
----------------------------------------------------------------------------
AYSYMMETRICAL FOOT TIMING
----------------------------------------------------------------------------
5. INTIAL CONTACT
"""
ic\_timing\_asymmetric\_total = id\_asymmetric\_timing\_error(
Trial\_ID, input\_df\_r, input\_df\_l
)
"""
----------------------------------------------------------------------------
AYSYMMETRICAL FOOT LANDING (Toe-Heel / Heel-Toe)
----------------------------------------------------------------------------
6. INITIAL CONTACT
"""
ic\_landing\_asymmetric\_total = asymmetric\_landing\_error(
Trial\_ID, ankle\_PF\_right, ankle\_PF\_left
)
"""
----------------------------------------------------------------------------
LATERAL TRUNK FLEXION
----------------------------------------------------------------------------
7. INITIAL CONTACT
"""
ic\_trunk\_lateral\_total = id\_lateral\_trunk\_flexion\_error(Trial\_ID, input\_df\_r)
"""
----------------------------------------------------------------------------
KNEE VALGUS
----------------------------------------------------------------------------
8. INITIAL CONTACT
16. MAXIMUM POSITION
"""
ic\_knee\_valgus\_total, max\_knee\_valgus\_total = id\_knee\_valgus\_error(
Trial\_ID, input\_df\_r, input\_df\_l
)
"""
----------------------------------------------------------------------------
WIDE STANCE WIDTH
----------------------------------------------------------------------------
9. INITIAL CONTACT
"""
ic\_hip\_wide\_total, ic\_hip\_add\_deg\_r, ic\_hip\_add\_deg\_l = id\_wide\_stance\_error(
Trial\_ID, input\_df\_r, input\_df\_l
)
"""
----------------------------------------------------------------------------
NARROW STANCE WIDTH
----------------------------------------------------------------------------
10. INITIAL CONTACT
"""
ic\_hip\_narrow\_total = id\_narrow\_stance\_error(
Trial\_ID, ic\_hip\_add\_deg\_r, ic\_hip\_add\_deg\_l
)
"""
----------------------------------------------------------------------------
FOOT INWARD ROTATION
----------------------------------------------------------------------------
11. INITIAL CONTACT
"""
ic\_foot\_inward\_total = id\_foot\_inward\_error(Trial\_ID, input\_df\_r, input\_df\_l)
"""
----------------------------------------------------------------------------
FOOT OUTWARD ROTATION
----------------------------------------------------------------------------
12. INITIAL CONTACT
"""
ic\_foot\_outward\_total = id\_foot\_outward\_error(Trial\_ID, input\_df\_r, input\_df\_l)
"""
----------------------------------------------------------------------------
ASYMMETRIC LOADING / HIP SHIFT
----------------------------------------------------------------------------
17. MAXIMUM POSITION
"""
max\_asymmetric\_loading\_total = id\_asymmetric\_loading\_error(
Trial\_ID, input\_df\_r, input\_df\_l
)
"""
----------------------------------------------------------------------------
SAGITTAL PLANE JOINT DISPLACEMENT
----------------------------------------------------------------------------
18. MAXIMUM POSITIONS
"""
joint\_displacement\_total = id\_joint\_displacement\_error(
Trial\_ID,
max\_hip\_flexion\_total,
max\_trunk\_flexion\_total,
max\_knee\_flexion\_total,
max\_knee\_flexion\_deg\_r,
max\_knee\_flexion\_deg\_l,
max\_hip\_flexion\_deg\_r,
max\_hip\_flexion\_deg\_l,
)
"""
----------------------------------------------------------------------------
OVERALL IMPRESSION
----------------------------------------------------------------------------
19. MAXIMUM POSITIONS
"""
overall\_impression\_total = id\_overall\_impression\_error(
Trial\_ID,
max\_hip\_flexion\_total,
max\_trunk\_flexion\_total,
max\_knee\_flexion\_total,
max\_knee\_valgus\_total,
ic\_knee\_valgus\_total,
)
"""
----------------------------------------------------------------------------
TOTAL SCORE
----------------------------------------------------------------------------
"""
LESS\_total = (
ic\_knee\_flexion\_total
+ max\_knee\_flexion\_total
+ ic\_knee\_valgus\_total
+ max\_knee\_valgus\_total
+ ic\_trunk\_lateral\_total
+ ic\_trunk\_flexion\_total
+ max\_trunk\_flexion\_total
+ ic\_hip\_flexion\_total
+ max\_hip\_flexion\_total
+ ic\_hip\_wide\_total
+ ic\_hip\_narrow\_total
+ ic\_foot\_inward\_total
+ ic\_foot\_outward\_total
+ ic\_ankle\_PF\_total
+ ic\_timing\_asymmetric\_total
+ ic\_landing\_asymmetric\_total
+ max\_asymmetric\_loading\_total
+ joint\_displacement\_total
+ overall\_impression\_total
)
LESS\_results.at[Trial\_ID, "Total\_Score"] = LESS\_total
LESS\_results\_short.at[Trial\_ID, "Total\_Score"] = LESS\_total
print(f"LESS\_total value in row '{Trial\_ID}': {LESS\_total}")
nan\_count = LESS\_results.loc[Trial\_ID].isnull().sum()
### uncomment to check if missing values present
print(f"Number of NaNs in row '{Trial\_ID}': {nan\_count}")
return LESS\_results
"""
-----------------------------------------------------------------------
---------------------------- Sub Functions ----------------------------
-----------------------------------------------------------------------
"""
# 1 - - - - - - - - - - - - - - - - - - - - - - - - - - - - - - - - - - - - - - -
def id\_knee\_flexion\_error(Trial\_ID, input\_df\_r, input\_df\_l):
"""
----------------------------------------------------------------------------
KNEE FLEXION
1. INITIAL CONTACT
13. MAXIMUM POSITION
Inputs: knee\_flexion\_r and knee\_flexion\_l from input df
Outputs: ERROR == IC value < 30 degrees
ERROR == Max value < 65 degrees
Notes: Greater POSITIVE values represent INCREASED Flexion
"""
ic\_error = 30.0
max\_error = 65.0
ic\_knee\_flexion\_deg\_r = input\_df\_r.loc[0, "knee\_angle\_r"]
max\_knee\_flexion\_deg\_r = input\_df\_r.loc[
0 : input\_df\_r["knee\_angle\_r"].idxmax(), "knee\_angle\_r"
].max()
ic\_knee\_flexion\_deg\_l = input\_df\_l.loc[0, "knee\_angle\_l"]
max\_knee\_flexion\_deg\_l = input\_df\_l.loc[
0 : input\_df\_l["knee\_angle\_l"].idxmax(), "knee\_angle\_l"
].max()
ic\_knee\_flexion\_total, max\_knee\_flexion\_total = score\_knee\_flexion(
ic\_error,
max\_error,
Trial\_ID,
ic\_knee\_flexion\_deg\_r,
max\_knee\_flexion\_deg\_r,
ic\_knee\_flexion\_deg\_l,
max\_knee\_flexion\_deg\_l,
)
LESS\_results.at[Trial\_ID, "1\_Knee\_Flexion\_IC\_R\_deg"] = ic\_knee\_flexion\_deg\_r
LESS\_results.at[Trial\_ID, "13\_Knee\_Flexion\_Displacement\_R\_deg"] = (
max\_knee\_flexion\_deg\_r
)
LESS\_results.at[Trial\_ID, "1\_Knee\_Flexion\_IC\_L\_deg"] = ic\_knee\_flexion\_deg\_l
LESS\_results.at[Trial\_ID, "13\_Knee\_Flexion\_Displacement\_L\_deg"] = (
max\_knee\_flexion\_deg\_l
)
return (
max\_knee\_flexion\_deg\_r,
max\_knee\_flexion\_deg\_l,
ic\_knee\_flexion\_total,
max\_knee\_flexion\_total,
)
def score\_knee\_flexion(
ic\_error,
max\_error,
Trial\_ID,
ic\_knee\_flexion\_deg\_r,
max\_knee\_flexion\_deg\_r,
ic\_knee\_flexion\_deg\_l,
max\_knee\_flexion\_deg\_l,
):
ic\_knee\_flexion\_r = 1 if ic\_knee\_flexion\_deg\_r < ic\_error else 0
max\_knee\_flexion\_r = 1 if max\_knee\_flexion\_deg\_r < max\_error else 0
ic\_knee\_flexion\_l = 1 if ic\_knee\_flexion\_deg\_l < ic\_error else 0
max\_knee\_flexion\_l = 1 if max\_knee\_flexion\_deg\_l < max\_error else 0
ic\_knee\_flexion\_total = 1 if ic\_knee\_flexion\_r or ic\_knee\_flexion\_l else 0
max\_knee\_flexion\_total = 1 if max\_knee\_flexion\_r or max\_knee\_flexion\_l else 0
LESS\_results.at[Trial\_ID, "1\_Knee\_Flexion\_IC\_R"] = ic\_knee\_flexion\_r
LESS\_results.at[Trial\_ID, "13\_Knee\_Flexion\_Displacement\_R"] = max\_knee\_flexion\_r
LESS\_results.at[Trial\_ID, "1\_Knee\_Flexion\_IC\_L"] = ic\_knee\_flexion\_l
LESS\_results.at[Trial\_ID, "13\_Knee\_Flexion\_Displacement\_L"] = max\_knee\_flexion\_l
LESS\_results.at[Trial\_ID, "1\_Knee\_Flexion\_IC\_T"] = ic\_knee\_flexion\_total
LESS\_results.at[Trial\_ID, "13\_Knee\_Flexion\_Displacement\_T"] = max\_knee\_flexion\_total
LESS\_results\_short.at[Trial\_ID, "1\_Knee\_Flexion\_IC\_T"] = ic\_knee\_flexion\_total
LESS\_results\_short.at[Trial\_ID, "13\_Knee\_Flexion\_Displacement\_T"] = (
max\_knee\_flexion\_total
)
return ic\_knee\_flexion\_total, max\_knee\_flexion\_total
# 2 - - - - - - - - - - - - - - - - - - - - - - - - - - - - - - - - - - - - - - -
def id\_hip\_flexion\_error(Trial\_ID, input\_df\_r, input\_df\_l):
"""
----------------------------------------------------------------------------
HIP FLEXION
----------------------------------------------------------------------------
2. INITIAL CONTACT
14. MAXIMUM POSITION
Inputs: hip\_flexion\_r and hip\_flexion\_l from input df
Outputs: ERROR == IC value < 30 degrees
ERROR == Max value < 45 degrees
Notes: Greater POSITIVE values represent INCREASED Flexion
"""
ic\_error = 30.0
max\_error = 45.0
ic\_hip\_flexion\_deg\_r = input\_df\_r.loc[0, "hip\_flexion\_r"]
max\_knee\_angle\_row\_r = input\_df\_r.loc[input\_df\_r["knee\_angle\_r"].idxmax()]
max\_hip\_flexion\_deg\_r = max\_knee\_angle\_row\_r["hip\_flexion\_r"]
ic\_hip\_flexion\_deg\_l = input\_df\_l.loc[0, "hip\_flexion\_l"]
max\_knee\_angle\_row\_l = input\_df\_l.loc[input\_df\_l["knee\_angle\_l"].idxmax()]
max\_hip\_flexion\_deg\_l = max\_knee\_angle\_row\_l["hip\_flexion\_l"]
ic\_hip\_flexion\_total, max\_hip\_flexion\_total = score\_hip\_flexion(
ic\_error,
max\_error,
Trial\_ID,
ic\_hip\_flexion\_deg\_r,
max\_hip\_flexion\_deg\_r,
ic\_hip\_flexion\_deg\_l,
max\_hip\_flexion\_deg\_l,
)
LESS\_results.at[Trial\_ID, "2\_Hip\_Flexion\_IC\_R\_deg"] = ic\_hip\_flexion\_deg\_r
LESS\_results.at[Trial\_ID, "14\_Hip\_Flexion\_Displacement\_R\_deg"] = (
max\_hip\_flexion\_deg\_r
)
LESS\_results.at[Trial\_ID, "2\_Hip\_Flexion\_IC\_L\_deg"] = ic\_hip\_flexion\_deg\_l
LESS\_results.at[Trial\_ID, "14\_Hip\_Flexion\_Displacement\_L\_deg"] = (
max\_hip\_flexion\_deg\_l
)
return (
max\_hip\_flexion\_deg\_r,
max\_hip\_flexion\_deg\_l,
ic\_hip\_flexion\_total,
max\_hip\_flexion\_total,
)
def score\_hip\_flexion(
ic\_error,
max\_error,
Trial\_ID,
ic\_hip\_flexion\_deg\_r,
max\_hip\_flexion\_deg\_r,
ic\_hip\_flexion\_deg\_l,
max\_hip\_flexion\_deg\_l,
):
ic\_hip\_flexion\_r = 1 if ic\_hip\_flexion\_deg\_r < ic\_error else 0
max\_hip\_flexion\_r = 1 if max\_hip\_flexion\_deg\_r < max\_error else 0
ic\_hip\_flexion\_l = 1 if ic\_hip\_flexion\_deg\_l < ic\_error else 0
max\_hip\_flexion\_l = 1 if max\_hip\_flexion\_deg\_l < max\_error else 0
ic\_hip\_flexion\_total = 1 if ic\_hip\_flexion\_r or ic\_hip\_flexion\_l else 0
max\_hip\_flexion\_total = 1 if max\_hip\_flexion\_r or max\_hip\_flexion\_l else 0
### Save each side and total
LESS\_results.at[Trial\_ID, "2\_Hip\_Flexion\_IC\_R"] = ic\_hip\_flexion\_r
LESS\_results.at[Trial\_ID, "14\_Hip\_Flexion\_Displacement\_R"] = max\_hip\_flexion\_r
LESS\_results.at[Trial\_ID, "2\_Hip\_Flexion\_IC\_L"] = ic\_hip\_flexion\_l
LESS\_results.at[Trial\_ID, "14\_Hip\_Flexion\_Displacement\_L"] = max\_hip\_flexion\_l
LESS\_results.at[Trial\_ID, "2\_Hip\_Flexion\_IC\_T"] = ic\_hip\_flexion\_total
LESS\_results.at[Trial\_ID, "14\_Hip\_Flexion\_Displacement\_T"] = max\_hip\_flexion\_total
LESS\_results\_short.at[Trial\_ID, "2\_Hip\_Flexion\_IC\_T"] = ic\_hip\_flexion\_total
LESS\_results\_short.at[Trial\_ID, "14\_Hip\_Flexion\_Displacement\_T"] = (
max\_hip\_flexion\_total
)
return ic\_hip\_flexion\_total, max\_hip\_flexion\_total
# 3 - - - - - - - - - - - - - - - - - - - - - - - - - - - - - - - - - - - - - - -
def id\_trunk\_flexion\_error(Trial\_ID, input\_df\_r):
"""
----------------------------------------------------------------------------
TRUNK FLEXION
----------------------------------------------------------------------------
3. INITIAL CONTACT
15. MAXIMUM POSITION
Inputs: lumbar\_extension from input right df
Outputs: ERROR == IC value > -10 degrees
ERROR == Max value > displacement <10
--> operationally defined as 55 (prior -30)
Notes: Trunk flexion will be computed from the opencap lumbar\_extension column
Greater POSITVE values represent EXTENSION
Greater NEGATIVE values represent FLEXION
"""
ic\_error = -10.0
max\_error = 10.0
ic\_trunk\_flexion\_deg = input\_df\_r.loc[0, "lumbar\_extension"]
max\_knee\_angle\_row = input\_df\_r.loc[input\_df\_r["knee\_angle\_r"].idxmax()]
max\_trunk\_flexion\_deg = max\_knee\_angle\_row["lumbar\_extension"]
ic\_trunk\_flexion\_total, max\_trunk\_flexion\_total = score\_trunk\_flexion(
ic\_error, max\_error, Trial\_ID, ic\_trunk\_flexion\_deg, max\_trunk\_flexion\_deg
)
LESS\_results.at[Trial\_ID, "3\_Trunk\_Flexion\_IC\_deg"] = ic\_trunk\_flexion\_deg
LESS\_results.at[Trial\_ID, "15\_Trunk\_Flexion\_Max\_deg"] = max\_trunk\_flexion\_deg
return (ic\_trunk\_flexion\_total, max\_trunk\_flexion\_total)
def score\_trunk\_flexion(
ic\_error, max\_error, Trial\_ID, ic\_trunk\_flexion\_deg, max\_trunk\_flexion\_deg
):
ic\_trunk\_flexion\_total = 1 if ic\_trunk\_flexion\_deg > ic\_error else 0
max\_trunk\_flexion\_total = (
1 if (ic\_trunk\_flexion\_deg - max\_trunk\_flexion\_deg) < max\_error else 0
)
LESS\_results.at[Trial\_ID, "3\_Trunk\_Flexion\_IC\_T"] = ic\_trunk\_flexion\_total
LESS\_results.at[Trial\_ID, "15\_Trunk\_Flexion\_Max\_T"] = max\_trunk\_flexion\_total
LESS\_results\_short.at[Trial\_ID, "3\_Trunk\_Flexion\_IC\_T"] = ic\_trunk\_flexion\_total
LESS\_results\_short.at[Trial\_ID, "15\_Trunk\_Flexion\_Max\_T"] = max\_trunk\_flexion\_total
return ic\_trunk\_flexion\_total, max\_trunk\_flexion\_total
# 4 - - - - - - - - - - - - - - - - - - - - - - - - - - - - - - - - - - - - - - -
def id\_ankle\_plantarflexion\_error(
Trial\_ID, coord\_file, sampling\_freq, coord\_cutoff\_freq
):
"""
----------------------------------------------------------------------------
ANKLE PLANTARFLEXION
----------------------------------------------------------------------------
4. INITIAL CONTACT
Inputs: left/right toes & heel joint coordinates from input coordinates df
Outputs: ERROR == IC value >=1 foot landed heel first
Notes: Reads in coordinate data, filters specific coordinates
ID's Initial Contact time for toes and heel
Determines which came first
"""
(
data,
time,
y\_coords\_left\_toes,
y\_coords\_left\_heel,
y\_coords\_right\_toes,
y\_coords\_right\_heel,
) = load\_coord\_file(coord\_file)
left\_toes\_filtered, left\_heel\_filtered, right\_toes\_filtered, right\_heel\_filtered = (
apply\_butterworth\_filter\_coords(
y\_coords\_left\_toes,
y\_coords\_left\_heel,
y\_coords\_right\_toes,
y\_coords\_right\_heel,
sampling\_freq,
coord\_cutoff\_freq,
)
)
"""
Left Side
"""
toes\_first\_minima\_time, toes\_first\_minima\_y\_coord, lt\_first\_minima\_index = (
id\_initialcontact\_toes(left\_toes\_filtered, time)
)
heel\_first\_minima\_time, lh\_first\_minima\_index = id\_initialcontact\_heels(
left\_heel\_filtered, time
)
ankle\_PF\_left = 1 if (toes\_first\_minima\_time) >= (heel\_first\_minima\_time) else 0
LESS\_results.at[Trial\_ID, "4\_Ankle\_Plantar\_Flexion\_IC\_L"] = ankle\_PF\_left
"""
Right Side
"""
toes\_first\_minima\_time\_r, toes\_first\_minima\_y\_coord, rt\_first\_minima\_index = (
id\_initialcontact\_toes(right\_toes\_filtered, time)
)
heel\_first\_minima\_time\_r, rh\_first\_minima\_index = id\_initialcontact\_heels(
right\_heel\_filtered, time
)
ankle\_PF\_right = (
1 if (toes\_first\_minima\_time\_r) >= (heel\_first\_minima\_time\_r) else 0
)
LESS\_results.at[Trial\_ID, "4\_Ankle\_Plantar\_Flexion\_IC\_R"] = ankle\_PF\_right
"""
Total Score
"""
ic\_ankle\_PF\_total = 1 if ankle\_PF\_right or ankle\_PF\_left else 0
LESS\_results.at[Trial\_ID, "4\_Ankle\_Plantar\_Flexion\_IC\_T"] = ic\_ankle\_PF\_total
LESS\_results\_short.at[Trial\_ID, "4\_Ankle\_Plantar\_Flexion\_IC\_T"] = ic\_ankle\_PF\_total
return ankle\_PF\_left, ankle\_PF\_right, ic\_ankle\_PF\_total
# 5 - - - - - - - - - - - - - - - - - - - - - - - - - - - - - - - - - - - - - - -
def id\_asymmetric\_timing\_error(Trial\_ID, input\_df\_r, input\_df\_l):
"""
----------------------------------------------------------------------------
AYSYMMETRICAL FOOT TIMING
----------------------------------------------------------------------------
5. INTIAL CONTACT
Inputs: time from right and left df
Outputs: ERROR == when value is >0.034 difference (2 frames)
One foot lands 34ms before the other
"""
timing\_error = 0.034
ic\_timing\_r = input\_df\_r.loc[0, "time"]
ic\_timing\_l = input\_df\_l.loc[0, "time"]
ic\_timing\_asymmetric\_total = (
1 if abs(ic\_timing\_r - ic\_timing\_l) > timing\_error else 0
)
LESS\_results.at[Trial\_ID, "5\_Asymmetrical\_Timing"] = ic\_timing\_asymmetric\_total
LESS\_results\_short.at[Trial\_ID, "5\_Asymmetrical\_Timing"] = (
ic\_timing\_asymmetric\_total
)
return ic\_timing\_asymmetric\_total
# 6 - - - - - - - - - - - - - - - - - - - - - - - - - - - - - - - - - - - - - - -
def asymmetric\_landing\_error(Trial\_ID, ankle\_PF\_right, ankle\_PF\_left):
"""
----------------------------------------------------------------------------
AYSYMMETRICAL FOOT LANDING (Toe-Heel / Heel-Toe)
----------------------------------------------------------------------------
6. INITIAL CONTACT
Inputs: ankle\_plantar\_flexion Right and Left
Outputs: ERROR == IC one foot landed heel-toe and the other toe-heel
"""
if ankle\_PF\_right == ankle\_PF\_left:
ic\_landing\_asymmetric\_total = 0
else:
ic\_landing\_asymmetric\_total = 1
LESS\_results.at[Trial\_ID, "6\_Asymmetrical\_HeelToe\_ToeHeel"] = (
ic\_landing\_asymmetric\_total
)
LESS\_results\_short.at[Trial\_ID, "6\_Asymmetrical\_HeelToe\_ToeHeel"] = (
ic\_landing\_asymmetric\_total
)
return ic\_landing\_asymmetric\_total
# 7 - - - - - - - - - - - - - - - - - - - - - - - - - - - - - - - - - - - - - - -
def id\_lateral\_trunk\_flexion\_error(Trial\_ID, input\_df\_r):
"""
----------------------------------------------------------------------------
LATERAL TRUNK FLEXION
----------------------------------------------------------------------------
7. INITIAL CONTACT
Inputs: lumbar\_bending from right\_dataframe
Outputs: ERROR == when value is < -5 or > 5 degrees (i.e. 5 deg to either side)
Notes: Trunk lateral flexion will be computed from the opencap lumbar\_bending
Greater POSITIVE values represent LEFT lateral flexion
Greater NEGATIVE values represent RIGHT lateral flexion
"""
trunk\_lateral\_error = 5.0
### Extracts raw joint angles from input dataframes right and left
ic\_trunk\_lateral\_deg = input\_df\_r.loc[0, "lumbar\_bending"]
###LESS Scoring Trunk Flexion IC and Maximum###
### Calculate the boolean operators
ic\_trunk\_lateral\_r = 1 if ic\_trunk\_lateral\_deg > trunk\_lateral\_error else 0
ic\_trunk\_lateral\_l = 1 if ic\_trunk\_lateral\_deg < -(trunk\_lateral\_error) else 0
ic\_trunk\_lateral\_total = 1 if ic\_trunk\_lateral\_r or ic\_trunk\_lateral\_l else 0
##Setting Values into new table
### Set the values in the new row for trials
LESS\_results.at[Trial\_ID, "7\_Lateral\_Trunk\_Flexion\_IC\_deg"] = ic\_trunk\_lateral\_deg
LESS\_results.at[Trial\_ID, "7\_Lateral\_Trunk\_Flexion\_IC\_R"] = ic\_trunk\_lateral\_r
LESS\_results.at[Trial\_ID, "7\_Lateral\_Trunk\_Flexion\_IC\_L"] = ic\_trunk\_lateral\_l
LESS\_results.at[Trial\_ID, "7\_Lateral\_Trunk\_Flexion\_IC\_T"] = ic\_trunk\_lateral\_total
LESS\_results\_short.at[Trial\_ID, "7\_Lateral\_Trunk\_Flexion\_IC\_T"] = (
ic\_trunk\_lateral\_total
)
return ic\_trunk\_lateral\_total
# 8 - - - - - - - - - - - - - - - - - - - - - - - - - - - - - - - - - - - - - - -
def id\_knee\_valgus\_error(Trial\_ID, input\_df\_r, input\_df\_l):
"""
----------------------------------------------------------------------------
KNEE VALGUS
----------------------------------------------------------------------------
8. INITIAL CONTACT
16. MAXIMUM POSITION
Inputs: hip\_rotation\_r & \_l and hip\_adduction\_r & \_l
Outputs: ERROR == If any of the following conditions:
1. hip\_rot > 5 and hip\_add > -1
2. hip\_rot > 5 at IC or >7 at MAX
3. hip\_add > 0
Notes: knee valgus will be computed from the opencap hip\_rotation and hip\_adduction
Greater POSITIVE values represent INCREASED Valgus
"""
#nerfed in 5May2024
### Extracts raw joint angles from input dataframes right and left
ic\_hip\_rotation\_deg\_r = input\_df\_r.loc[0, "hip\_rotation\_r"]
max\_knee\_angle\_row\_r = input\_df\_r.loc[input\_df\_r["knee\_angle\_r"].idxmax()]
max\_hip\_rotation\_deg\_r = max\_knee\_angle\_row\_r["hip\_rotation\_r"]
ic\_hip\_rotation\_deg\_l = input\_df\_l.loc[0, "hip\_rotation\_l"]
max\_knee\_angle\_row\_l = input\_df\_l.loc[input\_df\_l["knee\_angle\_l"].idxmax()]
max\_hip\_rotation\_deg\_l = max\_knee\_angle\_row\_l["hip\_rotation\_l"]
ic\_hip\_adduction\_deg\_r = input\_df\_r.loc[0, "hip\_adduction\_r"]
max\_knee\_angle\_row\_r = input\_df\_r.loc[input\_df\_r["knee\_angle\_r"].idxmax()]
max\_hip\_adduction\_deg\_r = max\_knee\_angle\_row\_r["hip\_adduction\_r"]
ic\_hip\_adduction\_deg\_l = input\_df\_l.loc[0, "hip\_adduction\_l"]
max\_knee\_angle\_row\_l = input\_df\_l.loc[input\_df\_l["knee\_angle\_l"].idxmax()]
max\_hip\_adduction\_deg\_l = max\_knee\_angle\_row\_l["hip\_adduction\_l"]
ic\_knee\_valgus\_r = knee\_valgus\_boolean(
ic\_hip\_rotation\_deg\_r, ic\_hip\_adduction\_deg\_r
)
max\_knee\_valgus\_r = knee\_valgus\_boolean(
max\_hip\_rotation\_deg\_r, max\_hip\_adduction\_deg\_r
)
ic\_knee\_valgus\_l = knee\_valgus\_boolean(
ic\_hip\_rotation\_deg\_l, ic\_hip\_adduction\_deg\_l
)
max\_knee\_valgus\_l = knee\_valgus\_boolean(
max\_hip\_rotation\_deg\_l, max\_hip\_adduction\_deg\_l
)
### Total Score
ic\_knee\_valgus\_total = 1 if ic\_knee\_valgus\_r or ic\_knee\_valgus\_l else 0
max\_knee\_valgus\_total = 1 if max\_knee\_valgus\_r or max\_knee\_valgus\_l else 0
LESS\_results.at[Trial\_ID, "8\_Medial\_Knee\_IC\_R\_deg"] = ic\_hip\_rotation\_deg\_r
LESS\_results.at[Trial\_ID, "16\_Medial\_Knee\_Max\_R\_deg"] = max\_hip\_rotation\_deg\_r
LESS\_results.at[Trial\_ID, "8\_Medial\_Knee\_IC\_R"] = ic\_knee\_valgus\_r
LESS\_results.at[Trial\_ID, "16\_Medial\_Knee\_Max\_R"] = max\_knee\_valgus\_r
LESS\_results.at[Trial\_ID, "8\_Medial\_Knee\_IC\_L\_deg"] = ic\_hip\_rotation\_deg\_l
LESS\_results.at[Trial\_ID, "16\_Medial\_Knee\_Max\_L\_deg"] = max\_hip\_rotation\_deg\_l
LESS\_results.at[Trial\_ID, "8\_Medial\_Knee\_IC\_L"] = ic\_knee\_valgus\_l
LESS\_results.at[Trial\_ID, "16\_Medial\_Knee\_Max\_L"] = max\_knee\_valgus\_l
LESS\_results.at[Trial\_ID, "8\_Medial\_Knee\_IC\_T"] = ic\_knee\_valgus\_total
LESS\_results.at[Trial\_ID, "16\_Medial\_Knee\_Max\_T"] = max\_knee\_valgus\_total
LESS\_results\_short.at[Trial\_ID, "8\_Medial\_Knee\_IC\_T"] = ic\_knee\_valgus\_total
LESS\_results\_short.at[Trial\_ID, "16\_Medial\_Knee\_Max\_T"] = max\_knee\_valgus\_total
return ic\_knee\_valgus\_total, max\_knee\_valgus\_total
def knee\_valgus\_boolean(hip\_rotation\_deg, hip\_adduction\_deg):
"""
Outputs: ERROR == If any of the following conditions:
1. hip\_rot > 5 and hip\_add > -1
2. hip\_rot >7 at MAX
3. hip\_add > 0
"""
knee\_valgus = (
1
if hip\_rotation\_deg > 8.0
and hip\_adduction\_deg > -1
#or hip\_rotation\_deg > 10
or hip\_adduction\_deg > 1
else 0
)
return knee\_valgus
# 9 - - - - - - - - - - - - - - - - - - - - - - - - - - - - - - - - - - - - - - - - -
def id\_wide\_stance\_error(Trial\_ID, input\_df\_r, input\_df\_l):
"""
----------------------------------------------------------------------------
WIDE STANCE WIDTH
----------------------------------------------------------------------------
9. INITIAL CONTACT
Inputs: hip\_adduction\_ from opencap
Outputs: ERROR == IC value < -12 degrees
Notes: stance width will be computed from the opencap hip\_adduction
Greater POSITIVE values represent ADDUCTION
\*Greater NEGATIVE values represent ABDUCTION\*
"""
wide\_stance\_error = -12.0
ic\_hip\_add\_deg\_r = input\_df\_r.loc[0, "hip\_adduction\_r"]
ic\_hip\_add\_deg\_l = input\_df\_l.loc[0, "hip\_adduction\_l"]
ic\_hip\_wide\_r = 1 if ic\_hip\_add\_deg\_r < wide\_stance\_error else 0
ic\_hip\_wide\_l = 1 if ic\_hip\_add\_deg\_l < wide\_stance\_error else 0
ic\_hip\_wide\_total = 1 if ic\_hip\_wide\_r or ic\_hip\_wide\_l else 0
LESS\_results.at[Trial\_ID, "9\_Stance\_Hip\_abd\_IC\_R\_deg"] = ic\_hip\_add\_deg\_r
LESS\_results.at[Trial\_ID, "9\_Stance\_Wide\_IC\_R"] = ic\_hip\_wide\_r
LESS\_results.at[Trial\_ID, "9\_Stance\_Hip\_abd\_IC\_L\_deg"] = ic\_hip\_add\_deg\_l
LESS\_results.at[Trial\_ID, "9\_Stance\_Wide\_IC\_L"] = ic\_hip\_wide\_l
LESS\_results.at[Trial\_ID, "9\_Stance\_Wide\_IC\_T"] = ic\_hip\_wide\_total
LESS\_results\_short.at[Trial\_ID, "9\_Stance\_Wide\_IC\_T"] = ic\_hip\_wide\_total
return ic\_hip\_wide\_total, ic\_hip\_add\_deg\_r, ic\_hip\_add\_deg\_l
# 10 - - - - - - - - - - - - - - - - - - - - - - - - - - - - - - - - - - - - - - - - -
def id\_narrow\_stance\_error(Trial\_ID, ic\_hip\_add\_deg\_r, ic\_hip\_add\_deg\_l):
"""
----------------------------------------------------------------------------
NARROW STANCE WIDTH
----------------------------------------------------------------------------
10. INITIAL CONTACT
Inputs: hip\_adduction\_ from opencap
Outputs: ERROR == IC value > 0 degrees
Notes: stance width will be computed from the opencap hip\_adduction
Greater POSITIVE values represent ADDUCTION
\*Greater NEGATIVE values represent ABDUCTION
"""
narrow\_stance\_error = 0
ic\_hip\_narrow\_r = 1 if ic\_hip\_add\_deg\_r > narrow\_stance\_error else 0
ic\_hip\_narrow\_l = 1 if ic\_hip\_add\_deg\_l > narrow\_stance\_error else 0
ic\_hip\_narrow\_total = 1 if ic\_hip\_narrow\_r or ic\_hip\_narrow\_l else 0
LESS\_results.at[Trial\_ID, "10\_Stance\_Narrow\_IC\_R"] = ic\_hip\_narrow\_r
LESS\_results.at[Trial\_ID, "10\_Stance\_Narrow\_IC\_L"] = ic\_hip\_narrow\_l
LESS\_results.at[Trial\_ID, "10\_Stance\_Narrow\_IC\_T"] = ic\_hip\_narrow\_total
LESS\_results\_short.at[Trial\_ID, "10\_Stance\_Narrow\_IC\_T"] = ic\_hip\_narrow\_total
return ic\_hip\_narrow\_total
# 11 - - - - - - - - - - - - - - - - - - - - - - - - - - - - - - - - - - - - - - - - -
def id\_foot\_inward\_error(Trial\_ID, input\_df\_r, input\_df\_l):
"""
----------------------------------------------------------------------------
FOOT INWARD ROTATION
----------------------------------------------------------------------------
11. INITIAL CONTACT
Inputs: subtalar\_angle\_r and \_l from opencap
Outputs: ERROR == IC value > 10 degrees
Notes: Foot rotation will be computed from the opencap subtalar\_angle\_
\*Greater POSITIVE values represent INWARD rotation\*
Greater NEGATIVE values represent OUTWARD rotation
"""
inward\_error = 10.0
ic\_subtalar\_deg\_r = input\_df\_r.loc[0, "subtalar\_angle\_r"]
ic\_subtalar\_deg\_l = input\_df\_l.loc[0, "subtalar\_angle\_l"]
ic\_foot\_inward\_r = 1 if ic\_subtalar\_deg\_r > inward\_error else 0
ic\_foot\_inward\_l = 1 if ic\_subtalar\_deg\_l > inward\_error else 0
ic\_foot\_inward\_total = 1 if ic\_foot\_inward\_r or ic\_foot\_inward\_l else 0
LESS\_results.at[Trial\_ID, "11\_IR\_Foot\_IC\_R"] = ic\_foot\_inward\_r
LESS\_results.at[Trial\_ID, "11\_IR\_Foot\_IC\_R\_deg"] = ic\_subtalar\_deg\_r
LESS\_results.at[Trial\_ID, "11\_IR\_Foot\_IC\_L"] = ic\_foot\_inward\_l
LESS\_results.at[Trial\_ID, "11\_IR\_Foot\_IC\_L\_deg"] = ic\_subtalar\_deg\_l
LESS\_results.at[Trial\_ID, "11\_IR\_Foot\_IC\_T"] = ic\_foot\_inward\_total
LESS\_results\_short.at[Trial\_ID, "11\_IR\_Foot\_IC\_T"] = ic\_foot\_inward\_total
return ic\_foot\_inward\_total
# 12 - - - - - - - - - - - - - - - - - - - - - - - - - - - - - - - - - - - - - - - - -
def id\_foot\_outward\_error(Trial\_ID, input\_df\_r, input\_df\_l):
"""
----------------------------------------------------------------------------
FOOT OUTWARD ROTATION
----------------------------------------------------------------------------
12. INITIAL CONTACT
Inputs: subtalar\_angle\_r and \_l from opencap
Outputs: ERROR == IC value < -15 degrees
Notes: Foot rotation will be computed from the opencap subtalar\_angle\_
Greater POSITIVE values represent INWARD rotation
\*Greater NEGATIVE values represent OUTWARD rotation\*
"""
outward\_error = (-15.0) #nerfed 5May2024
ic\_subtalar\_deg\_r = input\_df\_r.loc[0, "subtalar\_angle\_r"]
ic\_subtalar\_deg\_l = input\_df\_l.loc[0, "subtalar\_angle\_l"]
ic\_foot\_outward\_r = 1 if ic\_subtalar\_deg\_r < outward\_error else 0
ic\_foot\_outward\_l = 1 if ic\_subtalar\_deg\_l < outward\_error else 0
ic\_foot\_outward\_total = 1 if ic\_foot\_outward\_r or ic\_foot\_outward\_l else 0
LESS\_results.at[Trial\_ID, "12\_ER\_Foot\_IC\_R"] = ic\_foot\_outward\_r
LESS\_results.at[Trial\_ID, "12\_ER\_Foot\_IC\_R\_deg"] = ic\_subtalar\_deg\_r
LESS\_results.at[Trial\_ID, "12\_ER\_Foot\_IC\_L"] = ic\_foot\_outward\_l
LESS\_results.at[Trial\_ID, "12\_ER\_Foot\_IC\_L\_deg"] = ic\_subtalar\_deg\_l
LESS\_results.at[Trial\_ID, "12\_ER\_Foot\_IC\_T"] = ic\_foot\_outward\_total
LESS\_results\_short.at[Trial\_ID, "12\_ER\_Foot\_IC\_T"] = ic\_foot\_outward\_total
return ic\_foot\_outward\_total
# 13 - - - - - - - - - - - - - - - - - - - - - - - - - - - - - - - - - - - - - - - - -
def id\_asymmetric\_loading\_error(Trial\_ID, input\_df\_r, input\_df\_l):
"""
----------------------------------------------------------------------------
ASYMMETRIC LOADING / HIP SHIFT
----------------------------------------------------------------------------
17. MAXIMUM POSITION
Inputs: hip\_adduction\_ from opencap
Outputs: ERROR == Max position if hip abduction angles at IC differ > 5 degrees
Notes: stance width will be computed from the opencap hip\_adduction
Greater POSITIVE values represent ADDUCTION
Greater NEGATIVE values represent ABDUCTION
"""
loading\_error = 5
max\_knee\_angle\_row\_r = input\_df\_r.loc[input\_df\_r["knee\_angle\_r"].idxmax()]
max\_hip\_abd\_deg\_r = max\_knee\_angle\_row\_r["hip\_adduction\_r"]
max\_knee\_angle\_row\_l = input\_df\_l.loc[input\_df\_l["knee\_angle\_l"].idxmax()]
max\_hip\_abd\_deg\_l = max\_knee\_angle\_row\_l["hip\_adduction\_l"]
max\_asymmetric\_loading\_total = (
1 if abs(max\_hip\_abd\_deg\_r - max\_hip\_abd\_deg\_l) > loading\_error else 0
)
LESS\_results.at[Trial\_ID, "17\_Asymmetrical\_Load\_R\_deg"] = max\_hip\_abd\_deg\_r
LESS\_results.at[Trial\_ID, "17\_Asymmetrical\_Load\_L\_deg"] = max\_hip\_abd\_deg\_l
LESS\_results.at[Trial\_ID, "17\_Asymmetrical\_Load\_T"] = max\_asymmetric\_loading\_total
LESS\_results\_short.at[Trial\_ID, "17\_Asymmetrical\_Load\_T"] = (
max\_asymmetric\_loading\_total
)
return max\_asymmetric\_loading\_total
# 14 - - - - - - - - - - - - - - - - - - - - - - - - - - - - - - - - - - - - - - - - -
def id\_joint\_displacement\_error(
Trial\_ID,
max\_hip\_flexion\_total,
max\_trunk\_flexion\_total,
max\_knee\_flexion\_total,
max\_knee\_flexion\_deg\_r,
max\_knee\_flexion\_deg\_l,
max\_hip\_flexion\_deg\_r,
max\_hip\_flexion\_deg\_l,
):
"""
----------------------------------------------------------------------------
SAGITTAL PLANE JOINT DISPLACEMENT
----------------------------------------------------------------------------
18. MAXIMUM POSITIONS
Inputs: knee, hip, trunk LESS scores; left and right knee and hip degrees
Outputs: joint\_displacement\_total
If max\_hip\_flexion\_total, max\_trunk\_flexion\_total, and max\_knee\_flexion\_total
are all equal to 0, and if the specified conditions for max\_knee\_flexion\_deg\_r,
max\_knee\_flexion\_deg\_l, max\_hip\_flexion\_deg\_r, and max\_hip\_flexion\_deg\_l
are met, the joint\_displacement\_total remains 0.
knee flexion >= 55
hip flexion >=73
Otherwise, if any of the conditions are not met, it becomes 1.
Finally, if the initial conditions are not satisfied (i.e., at least one
of max\_hip\_flexion\_total, max\_trunk\_flexion\_total, and
max\_knee\_flexion\_total is not equal to 0), joint\_displacement\_total is set
to 2.
"""
max\_knee\_error = 55
max\_hip\_error = 73
if (
(max\_hip\_flexion\_total == 0)
and (max\_trunk\_flexion\_total == 0)
and (max\_knee\_flexion\_total == 0)
):
if (max\_knee\_flexion\_deg\_r >= max\_knee\_error) and (
max\_knee\_flexion\_deg\_l >= max\_knee\_error
):
if (max\_hip\_flexion\_deg\_r >= max\_hip\_error) and (
max\_hip\_flexion\_deg\_l >= max\_hip\_error
):
joint\_displacement\_total = 0
else:
joint\_displacement\_total = 1
else:
joint\_displacement\_total = 1
else:
joint\_displacement\_total = 2
LESS\_results.at[Trial\_ID, "18\_Joint\_Displacement\_T"] = joint\_displacement\_total
LESS\_results\_short.at[Trial\_ID, "18\_Joint\_Displacement\_T"] = (
joint\_displacement\_total
)
return joint\_displacement\_total
# 15 - - - - - - - - - - - - - - - - - - - - - - - - - - - - - - - - - - - - - - - - -
def id\_overall\_impression\_error(
Trial\_ID,
max\_hip\_flexion\_total,
max\_trunk\_flexion\_total,
max\_knee\_flexion\_total,
max\_knee\_valgus\_total,
ic\_knee\_valgus\_total,
):
"""
----------------------------------------------------------------------------
OVERALL IMPRESSION
----------------------------------------------------------------------------
19. MAXIMUM POSITIONS
Inputs: knee, hip, valgus LESS scores
Outputs: overall\_impression\_total
This code first checks if all the specified variables (max\_hip\_flexion\_total,
max\_trunk\_flexion\_total, max\_knee\_flexion\_total, and max\_knee\_valgus\_total)
are equal to 0. If they are all 0, overall\_impression\_total is
set to 0.
Next, it checks if either max\_hip\_flexion\_total or max\_knee\_flexion\_total is
equal to 1, and also if either max\_knee\_valgus\_total or ic\_knee\_valgus\_total
is equal to 1. If both conditions are true, overall\_impression\_total is
set to 2.
If none of the above conditions are met, overall\_impression\_total is
set to 1.
"""
if (
max\_hip\_flexion\_total == 0
and max\_trunk\_flexion\_total == 0
and max\_knee\_flexion\_total == 0
and max\_knee\_valgus\_total == 0
):
overall\_impression\_total = 0
elif (
max\_hip\_flexion\_total == 1
or max\_trunk\_flexion\_total == 1
or max\_knee\_flexion\_total == 1
) and (max\_knee\_valgus\_total == 1 or ic\_knee\_valgus\_total == 1):
overall\_impression\_total = 2
else:
overall\_impression\_total = 1
LESS\_results.at[Trial\_ID, "19\_Overall\_Impression\_T"] = overall\_impression\_total
LESS\_results\_short.at[Trial\_ID, "19\_Overall\_Impression\_T"] = (
overall\_impression\_total
)
return overall\_impression\_total
### %% [markdown]
### ### Cleaning Function
# %%
def bilateral\_process\_file(
file\_name, kinematics\_name, sampling\_freq, cutoff\_freq, coord\_cutoff\_freq
):
"""
Process the data from a .trc and .mot file and perform various operations.
1. ID's coordinates for L/R Big toe during movement (following filtering)
2. ID's first inverse peak of big toe coordinate (initial contact)
3. ID's last inverse peak prior vertical jump (toe off)
4. Applies the event times to the .mot kinematics file for L/R limbs
5. Applies 4th order butterworth to kinematic files
Args:
file\_name (str): File path of the .trc file.
kinematics\_name (str): File path of the kinematics data file.
Returns:
Stance phase of the DVJ for R and L LE joint kinematics
left\_kinematics\_{file\_name}.csv
right\_stance\_kinematics\_{file\_name}.csv
left\_kin\_{file\_name}\_filtered.csv
right\_kin\_{file\_name}\_filtered.csv
"""
title = os.path.splitext(os.path.basename(file\_name))[0]
print("Processing data from:", title)
### Load the .trc file
(
data,
time,
y\_coords\_left\_toes,
y\_coords\_left\_heel,
y\_coords\_right\_toes,
y\_coords\_right\_heel,
) = load\_coord\_file(file\_name)
"""
Filter Coordinates
"""
left\_toes\_filtered, left\_heel\_filtered, right\_toes\_filtered, right\_heel\_filtered = (
apply\_butterworth\_filter\_coords(
y\_coords\_left\_toes,
y\_coords\_left\_heel,
y\_coords\_right\_toes,
y\_coords\_right\_heel,
sampling\_freq,
coord\_cutoff\_freq,
)
)
"""
LEFT SIDE PROCESSING
"""
### - - - - - - IDENTIFY INITIAL CONTACT TOES - - - - - - #
toes\_first\_minima\_time, toes\_first\_minima\_y\_coord, lt\_first\_minima\_index = (
id\_initialcontact\_toes(left\_toes\_filtered, time)
)
### - - - - - - INITIAL CONTACT HEEL - - - - - - #
heel\_first\_minima\_time, lh\_first\_minima\_index = id\_initialcontact\_heels(
left\_heel\_filtered, time
)
##### index IC is determined by which hit ground first
y\_min\_index\_l, y\_min\_index\_l\_time = compare\_initialcontact\_toe\_vs\_heel(
toes\_first\_minima\_time,
lt\_first\_minima\_index,
heel\_first\_minima\_time,
lh\_first\_minima\_index,
time,
)
### - - - - - - IDENTIFY TOE OFF - - - - - - #
toe\_off\_index\_l, toes\_second\_minima\_time = id\_takeoff\_toes(
left\_toes\_filtered, time, lt\_first\_minima\_index
)
### - - - - - - Left: Plot the results - - - - - - - #
generate\_plot\_table\_quality\_check(
time,
left\_heel\_filtered,
left\_toes\_filtered,
y\_min\_index\_l\_time,
toes\_second\_minima\_time,
title,
"left",
)
### - - - - Left: Clip motion and coordinate files to IC and TO - - - - -
left\_kinematics, l\_stance\_coords = apply\_stance\_phase\_to\_motion\_file(
data,
time,
y\_min\_index\_l,
toe\_off\_index\_l,
kinematics\_name,
start\_column=14, #these may need to be updated hip\_flexion\_l
end\_column=20,
)
"""
Filter Left kinematics
"""
l\_filtered\_data = apply\_butterworth\_filter(
left\_kinematics, sampling\_freq, cutoff\_freq
)
save\_filtered\_data(l\_filtered\_data, title, side\_of\_body="left")
"""
Right side processing
"""
### - - - - - - IDENTIFY INITIAL CONTACT TOES - - - - - - #
toes\_first\_minima\_time, toes\_first\_minima\_y\_coord, rt\_first\_minima\_index = (
id\_initialcontact\_toes(right\_toes\_filtered, time)
)
### - - - - - - INITIAL CONTACT HEEL - - - - - - #
heel\_first\_minima\_time, rh\_first\_minima\_index = id\_initialcontact\_heels(
right\_heel\_filtered, time
)
##### index IC is determined by which hit ground first
y\_min\_index\_r, y\_min\_index\_r\_time = compare\_initialcontact\_toe\_vs\_heel(
toes\_first\_minima\_time,
rt\_first\_minima\_index,
heel\_first\_minima\_time,
rh\_first\_minima\_index,
time,
)
### - - - - - - IDENTIFY TOE OFF - - - - - - #
toe\_off\_index\_r, toes\_second\_minima\_time = id\_takeoff\_toes(
right\_toes\_filtered, time, rt\_first\_minima\_index
)
### - - - - - Right: Plot the results - - - - - -
generate\_plot\_table\_quality\_check(
time,
right\_heel\_filtered,
right\_toes\_filtered,
y\_min\_index\_r\_time,
toes\_second\_minima\_time,
title,
"right",
)
### - - - - - - Right: Clip motion and coordinate files to IC and TO - - - - - -
right\_kinematics, r\_stance\_coords = apply\_stance\_phase\_to\_motion\_file(
data,
time,
y\_min\_index\_r,
toe\_off\_index\_r,
kinematics\_name,
start\_column=7,
end\_column=13,
)
"""
Filter and save right kinematics
"""
r\_filtered\_data = apply\_butterworth\_filter(
right\_kinematics, sampling\_freq, cutoff\_freq
)
save\_filtered\_data(r\_filtered\_data, title, side\_of\_body="right")
return (
l\_stance\_coords,
r\_stance\_coords,
left\_kinematics,
right\_kinematics,
)
"""
-----------------------------------------------------------------------
---------------------------- Sub Functions ----------------------------
-----------------------------------------------------------------------
"""
def id\_takeoff\_toes(toes\_filtered, time, first\_minima\_index):
"""
Identifies timepoint of toe takeoff from ground contact
"""
toes\_local\_minima\_indices = argrelmin(toes\_filtered)
toes\_next\_peak\_index = (
np.argmax(toes\_filtered[first\_minima\_index:]) + first\_minima\_index
)
toes\_next\_peak\_time = time[toes\_next\_peak\_index]
toes\_next\_peak\_value = toes\_filtered[toes\_next\_peak\_index]
last\_minima\_index = toes\_local\_minima\_indices[0][
np.where(toes\_local\_minima\_indices[0] < toes\_next\_peak\_index)[0][-1]
]
toes\_second\_minima\_time = (
time[last\_minima\_index] if last\_minima\_index is not None else None
)
toes\_second\_minima\_y\_coord = (
toes\_filtered[last\_minima\_index] if last\_minima\_index is not None else None
)
toe\_off\_index = last\_minima\_index
return toe\_off\_index, toes\_second\_minima\_time
def compare\_initialcontact\_toe\_vs\_heel(
toes\_first\_minima\_time,
toe\_first\_minima\_index,
heel\_first\_minima\_time,
heel\_first\_minima\_index,
time,
):
"""
Determines whether the toes or heel landed first
"""
if toes\_first\_minima\_time <= heel\_first\_minima\_time:
y\_min\_index = toe\_first\_minima\_index
else:
y\_min\_index = heel\_first\_minima\_index
y\_min\_index = y\_min\_index #Made Edits HERE! 15Apr -12 frames (.05 seconds)
y\_min\_index\_time = time[y\_min\_index]
return y\_min\_index, y\_min\_index\_time
def plot\_filtered\_data(
time,
heel\_filtered,
toes\_filtered,
y\_min\_index\_time,
toes\_second\_minima\_time,
title,
side,
):
fig, ax = plt.subplots(figsize=(10, 5))
ax.plot(time, heel\_filtered, label="Filtered Y-coordinates Heel")
ax.plot(time, toes\_filtered, label="Filtered Y-coordinates Toes")
ax.axvline(y\_min\_index\_time, color="red", linestyle="--", label="Initial Contact")
ax.axvline(toes\_second\_minima\_time, color="green", linestyle="--", label="Toe-off")
ax.set\_xlabel("Time (s)")
ax.set\_ylabel("Y-coordinate (m)")
ax.legend()
plt.title(title + f" {side}")
plt.show()
def generate\_plot\_table\_quality\_check(
time,
heel\_filtered,
toes\_filtered,
y\_min\_index\_time,
toes\_second\_minima\_time,
title,
side,
):
plot\_filtered\_data(
time,
heel\_filtered,
toes\_filtered,
y\_min\_index\_time,
toes\_second\_minima\_time,
title,
side,
)
table = PrettyTable()
table.field\_names = ["Event", "Time (s)"]
table.add\_row(["Time of {side} initial contact", "{:.3f}".format(y\_min\_index\_time)])
table.add\_row(["Time of {side} toe-off", "{:.3f}".format(toes\_second\_minima\_time)])
table.add\_row(
[
f"{side} stance time duration",
"{:.3f}".format(toes\_second\_minima\_time - y\_min\_index\_time),
]
)
table.title = f"Results for {side} {title}"
print(table)
def apply\_stance\_phase\_to\_motion\_file(
data,
time,
y\_min\_index,
toe\_off\_index,
kinematics\_name,
start\_column: int,
end\_column: int,
):
"""
Clips motion file to be from initial contact to toe-off
"""
### Starting column number for left side kinematics (excluding the time column)
### Ending column number for left side kinematics (excluding the time column)
stance\_coords = pd.DataFrame(data)
stance\_coords = stance\_coords[
(stance\_coords.iloc[:, 1] >= time[y\_min\_index])
& (stance\_coords.iloc[:, 1] <= time[toe\_off\_index])
]
stance\_coords.reset\_index(drop=True, inplace=True)
title = os.path.splitext(os.path.basename(kinematics\_name))[0]
print("Processing left side data from:", title)
df\_kinematics = pd.read\_csv(kinematics\_name, sep="\t", header=8)
stance\_kinematics = pd.DataFrame(df\_kinematics)
kinematics = stance\_kinematics.iloc[
:,
list(range(0, 7))
+ list(range(start\_column, end\_column + 1))
+ list(range(21, 25)),
]
kinematics = kinematics[
(kinematics.iloc[:, 0] >= time[y\_min\_index])
& (kinematics.iloc[:, 0] <= time[toe\_off\_index])
]
kinematics.reset\_index(drop=True, inplace=True)
return kinematics, stance\_coords
# - - - - - - - - - - - - - - - - - - - - - - - - - - - - - - - - - - - - - - -
def save\_filtered\_data(filtered\_data, title, side\_of\_body=str):
filtered\_file\_name = f"{side\_of\_body}\_kin\_{title}\_filtered.csv"
filtered\_file\_name = filtered\_file\_name.replace(".mot", "")
file\_path = filtered\_file\_name
filtered\_data.to\_csv(file\_path, index=False)
print("Filtered data saved to:", filtered\_file\_name)
return
### %% [markdown]
### ### Processing Function
# %%
def remove\_specific\_trials(trial\_list, trial\_names\_to\_remove):
"""
Removes named trials from the list if there's an error
"""
names\_to\_remove\_set = set(trial\_names\_to\_remove)
return [
trial
for trial in trial\_list
if not any(name in trial for name in names\_to\_remove\_set)
]
def organize\_motion\_files(folder, motion, LESS, trials\_to\_remove):
"""
Organizing motion files in selected directory
Inputs:
1. folder directory
2. motion files location -- default
3. LESS is the common naming convention across trials
4. Trials that couldn't process -- manual
"""
motion\_path = os.path.join(folder, motion)
motion\_files = []
motion\_trials = [x for x in os.listdir(motion\_path) if LESS in x]
print(motion\_trials)
motion\_trials = remove\_specific\_trials(motion\_trials, trials\_to\_remove)
print(motion\_trials)
motion\_files = [motion + "/" + x for x in motion\_trials]
total\_motion\_files = len(motion\_files)
print("Total number of files in motion\_files:", total\_motion\_files)
print()
print("Check Motion Names")
for item in motion\_files:
print(item)
return motion\_files, motion\_trials
def organize\_marker\_files(folder, marker, motion\_trials):
"""
Organizing marker files in selected directory
Inputs:
1. folder directory
2. marker files location -- default
3. names of the motion trials
"""
marker\_files = []
marker\_path = os.path.join(folder, marker)
for o in motion\_trials:
marker\_trials = [
x for x in os.listdir(marker\_path) if o[:-4] in x and "miss" not in x
]
if marker\_trials:
marker\_files.append(marker + "/" + marker\_trials[0])
print()
total\_marker\_files = len(marker\_files)
print("Total number of files in marker\_files:", total\_marker\_files)
print()
print("Check Marker Names")
print("Confirm Order is maintained between motion and marker files")
for item in marker\_files:
print(item)
return marker\_files
def organize\_right\_trial\_files():
"""
Identifies and organizes cleaned RIGHT kinematic .csv files
"""
right\_trial\_files = []
right\_trial\_files = [x for x in os.listdir() if "right\_kin\_" in x]
right\_trial\_files.sort() # Sort the list in ascending order
print()
total\_files = len(right\_trial\_files)
print("Total number of files in right kin files:", total\_files)
print()
for item in right\_trial\_files:
print(item)
print()
return right\_trial\_files
def organize\_left\_trial\_files():
"""
Identifies and organizes cleaned LEFT kinematic .csv files
"""
left\_trial\_files = []
left\_trial\_files = [x for x in os.listdir() if "left\_kin\_" in x]
left\_trial\_files.sort()
print()
total\_files = len(left\_trial\_files)
print("Total number of files in left kin files:", total\_files)
print()
for item in left\_trial\_files:
print(item)
print()
return left\_trial\_files
def organize\_coord\_files(folder, marker, LESS, trials\_to\_remove):
"""
Identifies and organizes RAW coordinate .trc files
removes specified trials
"""
coord\_path = os.path.join(folder, marker)
coord\_files = []
coord\_files = [x for x in os.listdir(coord\_path) if LESS in x]
coord\_files.sort()
coord\_files = remove\_specific\_trials(coord\_files, trials\_to\_remove)
coord\_files = [file for file in coord\_files if not any(trial in file for trial in trials\_to\_remove)]
coord\_files = [os.path.join(coord\_path, x) for x in coord\_files] #new line
#coord\_files = [coord\_path + "/" + x for x in coord\_files] old
print()
total\_files = len(coord\_files)
print("Total number of files in marker files:", total\_files)
print()
print("marker files")
for item in coord\_files:
print(item)
print()
return coord\_files
'''
##### Process Trials ###
#
#
### > Step 1: Run all above for the packages and functions to be read in
#
### > Step 2: Confirm location of .mot and .trc files, either in original OpenCap download folder or a new batch trial folder
Must keep .mot files in OpenSimData/Kinematics folder and .trc in MarkerData folder (or change the variable names below)
#
### > Step 3: Clip and Filter .mot files
#
### > Step 4: Score LESS using filtered files
#
### > Step 5: Export .csv with results
#
### > Optional: Clear/Reset Dataframe
'''
# %%
### Run this between subjects to easily append the same excel sheet
os.getcwd()
### %% [markdown]
### ## STEP 3: Clip and Filter
#
### 1. Name the folder within your drive
### 2. Fill in the naming convention for what was used for LESS recordings
### 3. Confirm that motion and marker are appropriately mapped within folder
### 4. When ready, run the code chunk
### 5. If any errors appear, copy the name of the trial error and remove them
#
### \*\*Reviewing Plots for Quality Control\*\*
### - \*If an answer is no. Remove the trial.\*
### 1. Does the plot follow a regular pattern?
### - The plot represents the vertical location in space of the person's foot
### - Starts above >.3m, then small peak, steep drop (landed), then 2nd peak
### - As long as the landing phase is captured, then it's okay (zone b/w peaks)
### 2. Are the lines for initial ground contact and toe off appropriate?
### - Initial contact: first instance of lowest point
### - Toe-off: last lowest point after initial contact (before 2nd peak)
#
#
### %% Run Cell for STEP 3
folder = "/filepath" # user input folderpath in drive
LESS = "LESS" # user input common anchor across trials e.g., "S1\_LESS\_T1"
motion = "OpenSimData/Kinematics" # default location for OpenCap .mot files
marker = "MarkerData" # default location for OpenCap .trc files
sampling\_freq = 240 # user input sampling frequency of opencap (60, 120, 240)
cutoff\_freq = 12 # user input butterworth lowpass filter for opencap motion .mot data
coord\_cutoff\_freq = (
12 # user input butterworth lowpass filter for opencap coordinate .trc data
)
### Define the trial names to remove (replace with the desired name or list of names)
### Reasons to remove: Caused an error, wrong placement of IC and TO, bad trial, duplicate
trials\_to\_remove = [
]
# -------------------------
### Set working directory to pull files from
### I recommend first making a new folder which contains all files of interest
os.chdir(folder)
"""
1. Motion Files
"""
motion\_files, motion\_trials = organize\_motion\_files(
folder, motion, LESS, trials\_to\_remove
)
"""
2. Marker Files
"""
marker\_files = organize\_marker\_files(folder, marker, motion\_trials)
### Process each file in the list
for marker\_data, motion\_data in zip(marker\_files, motion\_files):
bilateral\_process\_file(
marker\_data, motion\_data, sampling\_freq, cutoff\_freq, coord\_cutoff\_freq
)
unique\_trials = set(trials\_to\_remove)
number\_of\_trials = len(unique\_trials)
print(f"Number of trials removed: {number\_of\_trials}")
LESS = "LESS"
coord\_files = organize\_coord\_files(folder, marker, LESS, trials\_to\_remove)
right\_trial\_files = organize\_right\_trial\_files()
### %% [Run Cell]
### ## STEP 4: Automatically Score LESS
###
#folder = "Filepath" # user input folderpath in drive
LESS = "LESS" # user input common anchor across trials e.g., "S1\_LESS\_T1"
motion = "OpenSimData/Kinematics" # default
marker = "MarkerData" # default
sampling\_freq = 240 # user input sampling frequency of opencap (60, 120, 240)
cutoff\_freq = 12 # user input butterworth lowpass filter for opencap motion .mot data
coord\_cutoff\_freq = (
12 # user input butterworth lowpass filter for opencap coordinate .trc data
)
right\_trial\_files = organize\_right\_trial\_files()
left\_trial\_files = organize\_left\_trial\_files()
coord\_files = organize\_coord\_files(folder, marker, LESS, trials\_to\_remove)
### Right, Left, and Marker named trials NEED TO MATCH and in order
print("TOTAL LESS SCORES")
### Iterate through matched right motion, left, motion, coordinate files and update LESS\_results
for right\_trial\_file, left\_trial\_file, coord\_file in zip(
right\_trial\_files, left\_trial\_files, coord\_files
):
print(f"processing {coord\_file}")
LESS\_results = process\_bilateral\_LESS(
right\_trial\_file,
left\_trial\_file,
coord\_file,
LESS\_results,
sampling\_freq,
coord\_cutoff\_freq,
)
print()
### %% [markdown]
### ### Check Results
### %% Run Cell here for:
### Check the results from LESS - confirm trials ordered correctly
pd.set\_option("display.max\_columns", 73) # Set the maximum number of columns to display
### Assuming LESS\_results is your DataFrame
print(LESS\_results)
print(LESS\_results\_short)
# %%
### Export Comprehensive LESS Scores to CSV file
LESS\_results.to\_csv('FILENAME\_less\_scores\_final.csv', index=False) #rename file path
### Export Short version LESS Scores to CSV file
LESS\_results\_short.to\_csv('FILENAME\_less\_short\_final.csv', index=False) #rename file path
# %%
### Clear dataframe and clipped motion files -- use this if there's an error and you want to restart
LESS\_results.drop(LESS\_results.index, inplace=True)
LESS\_results\_short.drop(LESS\_results\_short.index, inplace=True)
### Clear the working directory
cwd = os.getcwd()
files = os.listdir(cwd)
for file in files:
if "kin" in file:
os.remove(os.path.join(cwd, file))
print(f"Removed file: {file}")
# %%
#clear LESS data frame only
LESS\_results.drop(LESS\_results.index, inplace=True)
LESS\_results\_short.drop(LESS\_results\_short.index, inplace=True)
LESS\_results
