## Supplemental File 2 for "Open Source, Open Science: Development of OpenLESS as the Automated Landing Error Scoring System"

**Angle	Distal Segment	Proximal Segment	Plane**

Trunk Flexion	Pelvis	Thigh	Sagittal

Hip Flexion	Thigh	Pelvis	Sagittal

Knee Flexion	Lower leg	Thigh	Sagittal

Ankle Flexion	Foot	Lower Leg	Sagittal

Lateral Trunk Flexion	Trunk	World	Frontal

Hip Adduction	Thigh	Pelvis	Frontal

Knee Valgus	Lower Leg	Thigh	Frontal

Trunk Rotation	Trunk	World	Transverse

Foot Rotation	Foot	World	Transverse

|  | **OPENCAP INPUTS** | **DEFINITIONS** |
| --- | --- | --- |
| **INITIAL CONTACT** | | |
| Knee Flexion | Knee angle | Less than 30˚ of knee flexion at initial contact |
| Hip Flexion | Hip flexion | Less than 30˚ of hip flexion at initial contact |
| Trunk Flexion | Lumbar extension^1^ | Less than 10˚ of trunk flexion at initial contact *(> -10˚)* ^1^ |
| Ankle PF | Toe Coordinates  Heel Coordinates | Heel contact at initial contact  Heel lands at same time or prior to toes |
| Asymmetrical Timing | Time | One foot lands at least ≥ 34 ms before the other initial contact |
| Asymmetrical  Heel-Toe | Ankle PF results | One foot lands heel-to-toe and the other lands toe-to-heel at initial contact  Ankle PF right ≠ left |
| Lateral Trunk Flexion | Lumbar bending | Greater than 5˚ lateral bending to either side |
| Knee Valgus | Hip rotation  Hip adduction^1^ | If any of the following conditions occur at initial contact:   - Hip rotation > 8˚ and hip adduction > -1˚ - OR - Hip adduction >1˚ |
| Wide Stance Width | Hip adduction^1^ | Greater than 12˚ of hip abduction at initial contact *(< -12˚)* ^1^ |
| Narrow Stance Width | Hip adduction^1^ | Greater than 0˚ of hip adduction at initial contact *(> 0˚)*^1^ |
| Foot Inward Rotation | Subtalar angle | Greater than 10˚ of foot rotation inward *(> 10˚)* |
| Foot Outward Rotation | Subtalar angle | Greater than 15 degrees of foot rotation outward *(< -15˚)* |
| **MAXIMUM POSITION** | | |
| Knee Flexion | Knee angle | Less than 65˚ of maximum knee flexion |
| Hip Flexion | Hip flexion | Less than 45˚ degrees of maximum hip flexion |
| Trunk Flexion | Lumbar extension | Less than 10˚ degrees of difference between initial contact and maximum position trunk flexion *(< 10˚)* ^1^ |
| Knee Valgus | Hip rotation  Hip adduction^1^ | If any of the following conditions occur at max position:   - Hip rotation > 8˚ and hip adduction > -1˚ - OR - Hip adduction >1˚ |
| Asymmetrical Loading | Hip adduction^1^ | Greater than 5.1 degree^2^ difference in maximum hip adduction between right and left legs |
| Sagittal Plane Joint Displacement | Max position knee, hip, and trunk results | Soft (0) = No errors for knee, hip, and trunk flexion max position  Average (1) = < 55˚ max knee flexion OR < 73˚ max hip flexion  Stiff (2) = Error present for knee, hip, or trunk flexion max position |
| **Overall Impression** | Max position knee, hip, trunk, valgus, and initial contact valgus results. | Excellent (0) = No errors for knee, hip, and trunk flexion max position AND no error for knee valgus max position  Average (1) = All others that do not class as Excellent or Poor  Poor (2) = Errors for knee flexion OR hip flexion OR trunk flexion max position AND error for either knee valgus at initial contact OR knee valgus max position |
| **TOTAL SCORE^3^** |  | Summation of all errors |
| ^1^ These values are inverted in OpenCap. The pipeline handles the OpenCap values in their original directionality.  ^2^ Mean absolute error for drop jump kinematics was 5.1˚ (2.3˚, 8.6˚). Ulrich et al. 2022.  ^3^ For averaging across repeated trials we recommend two approaches 1) simple approach would be to average the errors for each item across a minimum of 3 trials, or 2) an error within an item occurs if most trials for an individual input was an error (e.g., ≥2 had errors out of 3 trials with knee flexion initial contact would have a final score of 1 error). Both approaches appear to represent the construct of movement errors similarly according to Hanzlikova et al. 2020. | | |
