## Supplemental File 3 for "Open Source, Open Science: Development of OpenLESS as the Automated Landing Error Scoring System"

| **Supplemental File 3.** Jump Landing Key Event Detection with Markerless Motion Capture | | | | |
| --- | --- | --- | --- | --- |
|  |  |  | **ICC (2,1)** | |
| **Jump Landing Event Time** | **Method** | **Mean (SD)** | **Value** | **P value** |
| Initial Ground Contact (seconds) | Force Plate | 6.745 (4.475) | 0.999 | <.001 |
|  | Marker-based | 6.775 (4.511) |  |  |
| Toe-Off (seconds) | Force Plate | 7.365 (4.524) | 0.999 | <.001 |
|  | Marker-based | 7.277 (4.552) |  |  |
| Notes: 12 post-ACLR subjects (6 males and 6 females), 98 trials comparing force plate and OpenCap derived key event times. Anterior Cruciate Ligament Reconstruction, ACLR; Intraclass Correlation Coefficient, ICC; Standard Deviation, SD. | | | | |
